# Assessing Dengue Virus Importation Risks in Africa: A Climate and Travel-Based Model

**DOI:** 10.1101/2024.05.07.24306997

**Authors:** Jenicca Poongavanan, José Lourenço, Joseph L.-H. Tsui, Vittoria Colizza, Yajna Ramphal, Cheryl Baxter, Moritz U.G. Kraemer, Marcel Dunaiski, Tulio de Oliveira, Houriiyah Tegally

**Affiliations:** Centre for Epidemic Response and innovation (CERI), Stellenbosch University, Stellenbosch, South Africa; BioISI (Biosystems and Integrative Sciences Institute), University of Lisbon, Lisbon, Portugal; Universidade Católica Portuguesa, Medical School, Biomedical Research Center, Lisboa, Portugal; Department of Biology, University of Oxford, Oxford, UK; Sorbonne Université, INSERM, Institut Pierre Louis d’Epidémiologie et de Santé Publique (IPLESP), Paris, France; Department of Biology, Georgetown University, Washington, District of Columbia, USA; Pandemic Sciences Institute, University of Oxford, UK; Computer Science Division, Department of Mathematical Sciences, Stellenbosch University, Stellenbosch, South Africa; KwaZulu-Natal Research Innovation and Sequencing Platform (KRISP), University of KwaZulu-Natal, Durban, South Africa

**Author notes:** Corresponding authors: Houriiyah Tegally and Tulio de Oliveira.

**Keywords:** Arboviruses, Dengue, Introductions, Mobility, Africa, Risk Flow

## Abstract

**Background:** Dengue is a significant global public health concern that poses a threat to Africa. Particularly, African countries are at risk of viral introductions through air travel connectivity with areas of South America and Asia that experience frequent explosive outbreaks. Limited reporting and diagnostic capacity hinder a comprehensive assessment of continent-wide transmission dynamics and deployment of surveillance strategies in Africa. This study aimed to identify African airports at high risk of receiving dengue infected passengers from Asia, Latin America and other African countries with high dengue incidence.

**Methods:** The risk of dengue introduction into Africa from countries of high incidence in Africa, Latin America and within Africa was estimated based on origin-destination air travel flows and epidemic activity at origin. We produced a novel proxy for local dengue epidemic activity using a composite index of theoretical climate-driven transmission suitability and population density, which we used, along with travel information in a risk flow model, to estimate importation risk.

**Findings:** We find that countries in East Africa face higher estimated risk of importation from Asia and other East African countries, whereas for West African countries, larger risk of importation is estimated from within the region. Some countries with high risk of importation experience low local transmission suitability which likely hampers the chances that importations lead to local transmission and establishment. Conversely, Mauritius, Uganda, Ivory Coast, Senegal, and Kenya are identified as countries susceptible to dengue introductions during periods of persistent transmission suitability.

**Interpretation:** Our work improves data driven allocation of surveillance resources, in regions of Africa that are at high risk of dengue introduction and establishment, including from regional circulation. This will be critical in detecting and managing imported cases and can improve local response to dengue outbreaks.

**Funding:** Rockefeller Foundation, National Institute of Health, EDCTP3 and Horizon Europe Research and Innovation, World Bank Group, Medical Research Foundation, Wellcome Trust, Google.org, Oxford Martin School Pandemic Genomics programme, John Fell Fund.

**Research in context:** *Evidence before this study:* Despite the significant global burden of dengue virus, Africa remains relatively understudied due to limited reporting and diagnostic capabilities. We searched PubMed for articles in English published on and before May 6, 2024, that included “Dengue OR dengue”, “Africa”, and “importation OR imported”. Few studies have investigated the introduction of dengue into African countries. Limited evidence includes phylogeographic studies describing a potential introduction of dengue from Brazil into Angola in 2013 and evidence of multiple historical introductions of dengue from Asia to Africa over several years. Before our study, none had employed a modelling framework to investigate the continental risks of importing dengue via viremic travellers into African countries from other regions of high dengue incidence.

*Added value of this study:* This study provides a novel approach to assessing the risk of dengue importation into Africa, integrating climate-dependent transmission suitability and air travel data. By identifying high-risk regions and highlighting the complex interplay between travel patterns, population density, and climatic factors, our findings enhance the understanding of ongoing dengue dynamics in Africa. This information enables targeted allocation of surveillance resources, improving preparedness and response to potential dengue outbreaks in susceptible regions.

*Implications of all the available evidence:* The integration of transmission suitability as a proxy for local epidemic activity and air travel data into a risk flow metric provides valuable insights into the risk of dengue importation into African airports from high-incidence countries. These findings have implications for tailored surveillance and prevention strategies in high-risk regions, facilitating early detection and management of potential imported dengue outbreaks in Africa.

## Introduction

Dengue is an arthropod-borne virus with four main serotypes and is a member of the genus *Flavivirus* of the family *Flaviviridae* (1,2). It is thought to cause between 5.2 million and 390 million infections around the world every year, mostly transmitted by *Aedes aegypti* and *Ae. albopictus* mosquitoes (3,4). Approximately half of the global human population is estimated to currently live in areas that are environmentally suitable for dengue transmission (3,5,6). Dengue is now endemic in more than 100 countries, with reported outbreaks predominantly in Central America, South America, and Southeast Asia, and has recently established epidemic cycles in parts of Africa and North America (6–8).

Despite the widespread distribution of mosquito vectors in Africa (9) and favourable transmission conditions characterised by high temperatures and increased urbanisation (7,10,11), severe gaps exist in our understanding of the transmission intensities of the virus on the continent. Limited diagnostic capacity and reporting on dengue incidence make it difficult to assess the true burden of dengue in Africa (3,5). Similarity of symptoms with other febrile illnesses such as malaria also play a role in underdiagnosis (12,13). Consensus of evidence studies have reported that dengue transmission is endemic in 34 countries in the African region (14,15). It is therefore speculated that dengue is more widespread in Africa than previously thought. In 2023 alone, 16 countries in Africa reported large dengue outbreaks (16).

Risk assessment for dengue in Africa has to consider several factors. Given frequent explosive dengue outbreaks in South America and Asia, African countries are, for instance, at risk of constant viral introductions through air travel connectivity. Once introduced, dengue can theoretically spread rapidly in large parts of Africa due to the presence of suitable vectors, appropriate environmental conditions and limited population immunity. Many initial dengue infections are also prone to be asymptomatic, which can lead to underreporting and unnoticed spread (1). While primary dengue infections are rarely fatal, secondary infections with a different dengue virus serotype can be highly problematic, potentially causing severe dengue fever (17). Even in countries where dengue is thought to be endemic, the importation of dengue genetic variants either from high incidence South American or Asian countries can exacerbate the burden of disease, favour serotype diversity and seed new explosive outbreaks.

The potential for dengue importation into a given location via viremic travellers is a function of transmission occurring in the origin country and the volume of travellers from such country to the destination country of interest. It is well documented that increased international travel and trade have facilitated dengue’s global spread and mixing of viral serotypes and genotypes (10,6,18,19). The influence of human mobility on pathogen spread has previously been characterised e.g. for influenza (20,21), Zika (22) and more recently, and very extensively, for SARS-CoV-2 (23,24). Human mobility also plays a key role in shaping individuals’ interactions with disease-carrying vectors and consequently affecting viral transmission (25,26). Understanding the combination of factors driving dengue introduction and onward transmission is crucial for effective surveillance, prevention, and control in Africa in the absence of antiviral treatment, adequate diagnosis, and widespread vaccine cover. This study aimed to identify African airports at high risk of receiving dengue infected passengers from Asia, Latin America and other African countries with high dengue incidence. In doing so, it identifies potential high risk areas and optimal periods (at a monthly scale) during which enhanced disease surveillance could contribute more to local public health.

## Methods

### 2.1 Data

#### 2.1.1 Air Travel

The air travel flow data used in this study was obtained from the International Air Transport Association (IATA). It comprises the number of origin-destination passenger tickets and accounts for any connections at intermediate airports for the year 2019. We opted for the year 2019 to reflect a recent customary year of travel preceding disruptions due to the COVID-19 pandemic. The data comprises monthly passenger volumes from 14 high incidence countries outside of Africa and 18 countries within the African continent that reported dengue outbreak in the last 10 years (selection described further down) to 54 African countries, encompassing all commercial airports (n = 197) in both the source and destination regions.

#### 2.1.2 Transmission Suitability Estimate

We sourced spatio-temporal estimates of climate-based transmission suitability of dengue (referred to as index P) developed and made accessible by estimates from (7) Nakase et al. (2023; https://doi.org/10.6084/m9.figshare.21502614). The index used in this study quantifies the transmission capacity of a single adult female mosquito throughout its lifetime in a completely susceptible host population, incorporating factors such as infectious periods and oviposition. For a more detailed explanation and validation of the P index, see (7), which provides an in-depth analysis and evidence supporting model, including spatio-temporal validation with regional dengue data from Brazil and Thailand. The index P metric utilises local temperature and humidity time series (spatial pixel resolution used was 0.25°□×□0.25° (∼28□km2) as primary inputs, which enables its application to any location with available climate data. For more detailed discussions on how climatic factors like temperature and rainfall affect mosquito biology and dengue transmission, see e.g. (27–29).

We post-processed raw climate-based transmission suitability P estimates to define periods of *persistence suitability.* Since P measures the number of hosts a single infected female may transmit DENV to during its infectious lifespan, a threshold of P > 1 represents the potential for epidemic expansion (i.e. single infected mosquitoes would transmit to more than one host). Following this rationale, periods of persistence suitability are then defined as the months where P > 1.0.

#### 2.1.3 Population Density

Population density at a given location significantly influences dengue transmission due to its role in determining the availability of human hosts for mosquito biting and thus the virus. To consider population density, we extracted the population count of each district within countries from the Gridded population of the World, using administrative (level 1) boundary data from the GADM database (30). To calculate the population density of a province, we aggregated the population count values of grid cells intersecting the province boundary and divided by the respective areas

### 2.2 Transmission suitability as a proxy for case counts

To address the challenge of missing or insufficient dengue case data, we explored the possibility of using climate-based transmission suitability (index P) as a proxy. To assess the viability of replacing dengue case data with transmission suitability, we conducted a preliminary correlation test between transmission suitability and monthly dengue cases for countries in which monthly dengue case data was available (see supplementary figure S1, p.1). Moreover, using transmission suitability instead of case data would allow for a more fine scaled understanding of dengue activity in a country, surpassing the resolution typically available at the national level.

For this study, we considered countries outside of Africa with large outbreaks of dengue in 2019 as flagged by the European Centre for Disease Prevention and Control (ECDC) (accessed on 10th May 2023; https://www.ecdc.europa.eu/en/dengue). These were predominantly in Latin America and Asia. Monthly dengue case data from these countries were then extracted from public health organisations such as the Pan American Health Organisation (PAHO; www3.paho.org), from governmental health reports or bulletins and from statistical bureau websites (governmental reporting websites, WHO reports, etc). The inclusion criteria for countries of high dengue incidence used in this study was based on the availability of monthly case data in 2019 as it allowed for reliable testing of the transmission suitability proxy and use of travel data for the same year. This resulted in a final selection of 14 countries of high dengue incidence in Latin America and Asia, namely: Brazil, Bolivia, Peru, Sri Lanka, Vietnam, Malaysia, Columbia, Thailand, Nicaragua, India, Bangladesh, Cambodia, Singapore and Belize. We also extracted province-level case data where openly available. In the correlation coefficient computation we also incorporated data from two African countries, Burkina Faso and Mauritius, for which monthly data for the year 2019 were available.

We use the Spearman’s Rank correlation coefficient to examine the relationship between transmission suitability and monthly dengue case counts for 16 countries. Following the methodology used by (7), we computed correlation coefficients between case counts and transmission suitability with various month lags (month+0, month+1 and month +2). We found (see supplementary figure S1, p.1) a significant positive correlation between transmission suitability and dengue cases for most countries, with the highest association at a lag of one month (12 out of 16 countries), corroborating previous analyses (7). The strength of the association ranged from −0.204 to 0.954. For some countries (n = 3), we found a weak correlation which could be caused by several factors, such as: i) limited case data obscuring seasonal patterns, which results in poor alignment with the index, ii) the national-level index failing to accurately reflect the overall case distribution if cases are concentrated in specific regions. Correlation between case data and transmission suitability was also calculated at higher resolution (State level) for Brazil, and the strongest relationship was also found at a one month lag (see supplementary figure S2, p.2). Thus, for the rest of the presented analyses, transmission suitability with a one-month lag was used. Subsequently, we included 18 African countries that have experienced a dengue outbreak in the past decade as reported in the WHO Afro reports. The 18 countries were: Angola, Benin, Burkina Faso, Chad, Comoros, Ethiopia, Ghana, Guinea, Kenya, Mali, Mauritania, Mauritius, Niger, Nigeria, Senegal, Tanzania, Togo and Sao Tome and Principe.

While the transmission suitability index serves as a valuable indicator of dengue risk in a given region, its standalone consideration provides only an answer to whether climate is suitable for transmission. Additional factors, such as the presence of the host and vector, are crucial for a more accurate assessment. Specifically, *Aedes* mosquito density data, along with host density and various other environmental factors (e.g. precipitation, vector habitat distribution, etc) that influence the spatial distribution of vectors, remain relevant for a complete understanding of the biological system. The main reason why such actors are often left out of modelling approaches is the lack of sufficient data with adequate spatio-temporal resolution. In this study, advancing on others based solely on climate-based suitability for vectors or viral transmission, we address one extra factor by incorporating human population density alongside the transmission suitability index. Using Spearman’s Rank correlation coefficient, we also examine the correlation between dengue cases and the composite index at a provincial level (see supplementary figure S3, p.3). We thus define a composite index (*t*):

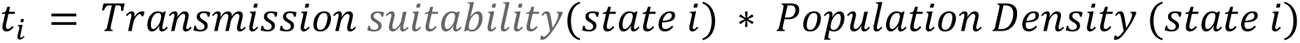

which captures the combined influence of the transmission suitability and the population density at the district level, by multiplying both elements (see supplementary figure S4 and S5, p.4).

### 2.3 Risk Flow Metric

The importation risk to each airport (destination) in each African country was estimated as the probability of importing a case from each state (origin) within each country of high incidence, accounting for the origin–destination travel flows and their different, estimated transmission suitability from the originating states. For cross-country comparisons, we aggregated the resulting risk of introduction from each airport to the country level, and for visualisation purposes, we calculated the average risk across each airport over the 12-month period (January - December 2019).

The methodology was adapted from (31) and is as follows:

Risk flow 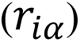 from origin state *i* to destination airport *α* is calculated by:

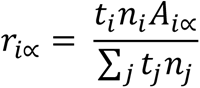

where *t_i_* is the combined transmission suitability and population density of state *i*, *n_i_* is the travel flux from the origin state *i*, *A_iα_* is the probability of a traveller flying from *i* to *α*, conditioned on travelling internationally from *i* (by construction, 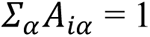). The denominator accounts for the transmission suitability in various states and the potential risk associated with travellers from different origin states. It is constructed as the sum product of all transmission suitability and travel flows originating from each individual state (denoted by j). This normalisation process ensures that the impact of each state’s transmission suitability and travel flow is proportionally represented in relation to the collective risk from all origin states.

The total risk of case importation to destination a is then given by

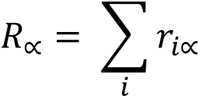

This risk is normalised such that 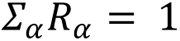.

To summarise and compare the overall introduction risks of dengue into African countries from high incidence countries in Asia and South America, we aggregated the risk originating from Asia, South America and Africa separately by summing it across all airports to the district level and then calculating the average value over all months. We then computed the proportions of risk coming from Asia, South America and Africa for each district. To further explore the dynamics between introduction risk and the local transmission suitability, we applied cosine similarity, a measure of synchrony between two time series. Cosine similarity helped us evaluate how closely the temporal patterns of introduction risk align with local transmission suitability. The cosine similarity ranges between 0 and 1, with 0 being less synchronous and 1 being highly synchronous.

Additionally, in aggregating the risk for destination countries, we conducted a secondary computation of risk emphasising the locations characterised by persistence suitability (see section 2.1.2). This was done by excluding any estimate of risk that occurred outside of times of persistence suitability (i.e. when index P <= 1) in respective destination locations. In other words, we filter only for estimated risks that are most likely to lead to onward transmission and local outbreaks. We term this introduction suitability while we term the risk based on the non-filtered values the raw introduction risk. It is important to note that the resulting risk values should not be interpreted in absolute terms but rather as indicators of the direction and timing of risk introduction, highlighting where and when the risk is coming in.

## Results

### Dengue introduction risks from high incidence Asian and South American countries into Africa

Several countries in Africa are estimated to have high transmission suitability for dengue and simultaneously high population density, indicating elevated potential for local transmission following viral introductions (Supplementary Figure S4, p.4). At the same time, certain countries on the continent are receiving variable, and sometimes high volumes of international passengers from various Asian and South American countries (Supplementary Figure S6, p.5). In 2019, over 504 million travellers entered the African continent from the 14 high-incidence countries. Of the total number of travellers, 39% originated from Southeast Asia, 34% from South Asia and 27% from South America (Supplementary Figure S6, p.5). The months with the highest travel volume entering the African continent from these places were November, December, and January.

The total risk flow of dengue from countries with high incidence rates, from Asia and South America showed that South Africa and Egypt were the countries at higher risk of dengue importation, generally considered as popular tourist destinations receiving large volumes of vacationers, followed by Kenya, Angola, Morocco, Seychelles and Mauritius (Figure 1). Egypt is exposed to significant risk primarily originating from Asia, specifically its biggest risk coming from Malaysia. Kenya, Mauritius, Tanzania, and Uganda exhibit an elevated risk of introduction from India. Risks from Singapore were also high, especially towards Mauritius, South Africa and Egypt. Risks from Vietnam are relatively low across the entire continent, with the most significant potential impact observed in South Africa and Angola. Additionally, Morocco and Nigeria are identified as regions with a considerable risk of disease introduction from Nicaragua.

**Figure 1.**
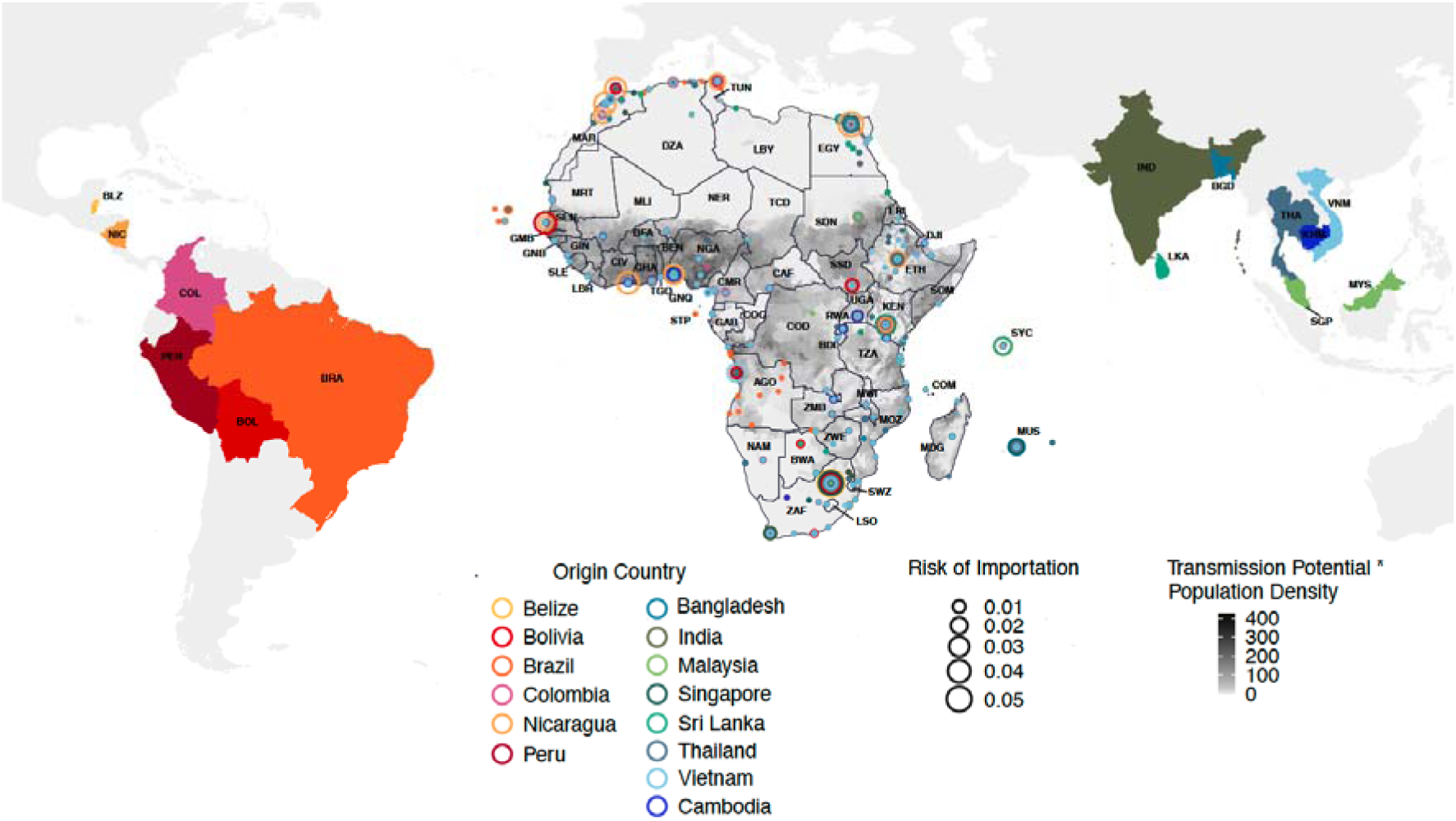
Mean risk of dengue introduction into African countries in 2019 from 14 countries in Asia and Latin America. The risk of dengue introduction into African countries from 14 origin countries in Asia and Latin America is represented by circles on the African continent. The size of the circles represent the size of the risk averaged over the 12 months for each airport in Africa. The colour of the circles represents the country from which the risk is coming from and the fill colour of those countries are consistently matched. The fill colour of the African continent represents the index of transmission suitability multiplied by population density to highlight the hotspots of high transmission at the destination.

Results revealed a general trend that countries in the southern and eastern African region are faced with higher risks of dengue importation from Asia, whereas central, western and a portion of northern African countries face higher risk of introduction from South America. South Africa is estimated to receive large introduction risks from both origin regions. When considering the raw risks of introductions overlayed onto the local transmission suitability index across Africa, it becomes clear that high risk of introduction is not necessarily linked to high transmission suitability (Figure 1). For instance, the high risks of introduction into South Africa may not necessarily translate to transmission locally due to very low dengue transmission suitability.

### Dengue introduction risks within Africa

The analysis of dengue introduction risk into African countries from other African countries with dengue circulation revealed significant regional patterns. We identified that countries in West Africa and East Africa exhibit higher importation risks, particular from regional transmission,compared to other regions (Figure 2). High importation risks are estimated into countries such as Kenya, Tanzania, Cameroon and Gabon. A notable pattern is that the importation risk within the East African region primarily originates from within the region itself, a trend also observed for West Africa. In contrast, countries in Northern Africa, such as Libya and Tunisia and those in Southern Africa, such as South Africa, exhibit lower dengue importation risks from the African continent itself.

**Figure 2:**
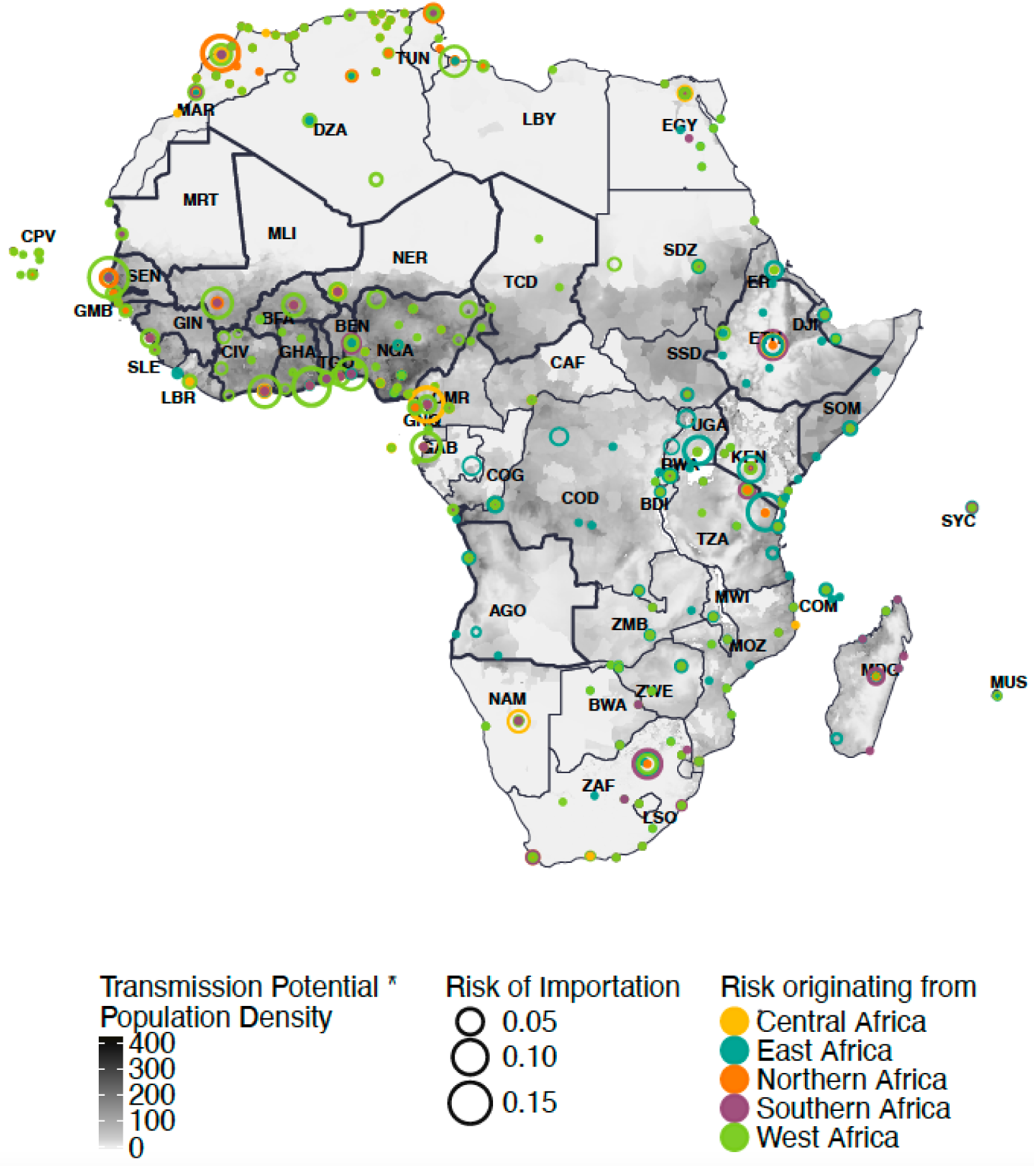
Mean risk of dengue introduction into African countries in 2019 from 18 African countries with Dengue circulation. Originating African countries considered here are Chad, Sao Tome and Principe (Central Africa), Comoros, Ethiopia, Kenya, Tanzania (East Africa), Benin, Burkina Faso, Ghana, Guinea, Mali, Niger, Nigeria, Senegal, Togo (West Africa), Angola, Mauritius (Southern Africa) and Mauritania (Northern Africa). The risk of dengue introduction into African countries is represented by circles on the African continent coloured by the corresponding region of the originating African country. The size of the circles represent the size of the risk averaged over the 12 months for each airport in Africa. The border for African countries considered as originating locations in the study are outlined in black. The fill colour of the African continent represents the index of transmission suitability multiplied by population density to highlight the hotspots of high transmission at the destination.

### Spatiotemporal variation in risks of introduction and local transmission suitability

In addition to quantifying the risk of dengue introductions from countries of high incidence in Asia and South America we evaluated the timing of these introductions and whether they would arrive at periods of high local transmission suitability in Africa. For this, the monthly introduction risk was mapped to local transmission suitability at the airport level in Africa. Egypt was estimated to be at high risk of dengue introduction during the second half of the year from both Asia and South America (Figure 3). This temporal pattern aligns with a concurrent period of heightened transmission suitability within the country, which could indicate a higher likelihood of viral introductions leading to onward transmission. To quantify the synchrony between the estimated viral importation risk and the local transmission suitability, we computed the cosine similarity (CS) between the different time-series, which revealed a CS of 0.311 for estimated risks from Asia and 0.147 from Latin America into Egypt (see Supplementary Table S1, p.10). In fact in 2023, Egypt experienced a large dengue outbreak around September to November (32), which coincides with peaks in estimated risk and transmission suitability. Angola and Ethiopia both were estimated to have a continuous risk of disease introduction from Asia throughout the year. However, in Angola, the transmission suitability was predicted to decline during the months of July, August, and September. Mauritius was also estimated to be at high risk of introduction from Asia. The cosine similarity between the risk of introduction and transmission suitability for Mauritius was 0.437 for Asia and 0.183 for South America. On the other hand, we observed high risks of dengue introduction into South Africa but, in general, the country exhibited a low transmission suitability throughout the year, indicating these potential introductions as likely impasses for transmission. In Nigeria, the majority of the high risk of introduction from South America were estimated to occur during periods of lower local transmission suitability. Angola and Mauritius were identified as being at high-risk of suitable introductions, given the combination of high and synchronous risk of introduction from Asia (see Supplementary Table S1, p.1).

**Figure 3.**
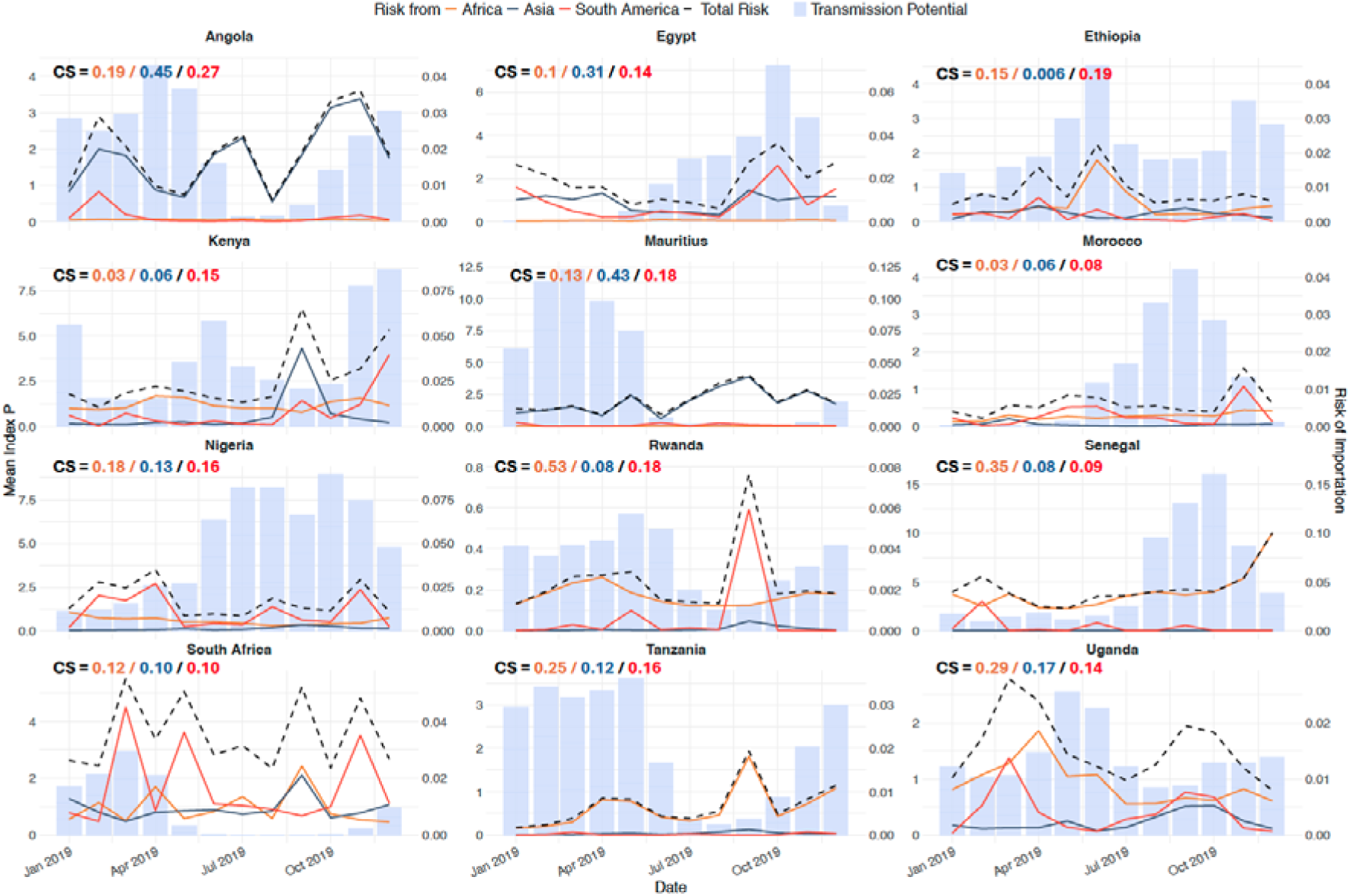
Time-varying risk of introduction into selected African countries in 2019. The blue, red and orange lines represent the risk of importation from Asia, South America and Africa, respectively, and the dotted line represents the total risk of importation from both continents and African countries across the year. The risk of importation was aggregated at the national level by summing the individual risks from all airports within each African destination country. The blue bars represent the time-varying transmission suitability index. The CS value represents the cosine similarity between the two timelines, where a value of 0 indicates low synchrony and a value of 1 indicates high synchrony. Here, we demonstrate countries that had the most synchronicity between transmission suitability and risk of introduction - see supplementary S7, p.6 for additional countries.

### Overall introduction suitability in Africa

We then integrated the risk of dengue importation from 14 countries outside of Africa with that from 18 African countries to gain a clearer understanding of the proportion of importation risk originating from within the African continent compared to that from external sources. We filtered for risk estimates that are most likely to lead to onward transmission: introduction suitability risk, that is, risks that occur during periods of persistence suitability (P>1). Figure 4A illustrates the aggregated raw risk of dengue introduction throughout the year 2019 across individual provinces or districts (administrative level 1) within each African country, and aggregated as originating either from Latin America, Asia, West, East, Central, Northern or Southern Africa. Conversely, Figure 4B presents the aggregated introduction suitability of dengue into Africa. Notably, in South Africa, where the transmission suitability (P) remains consistently below 1 throughout the year, the risk of dengue evolving into an outbreak from an introduction is negligible.

**Figure 4:**
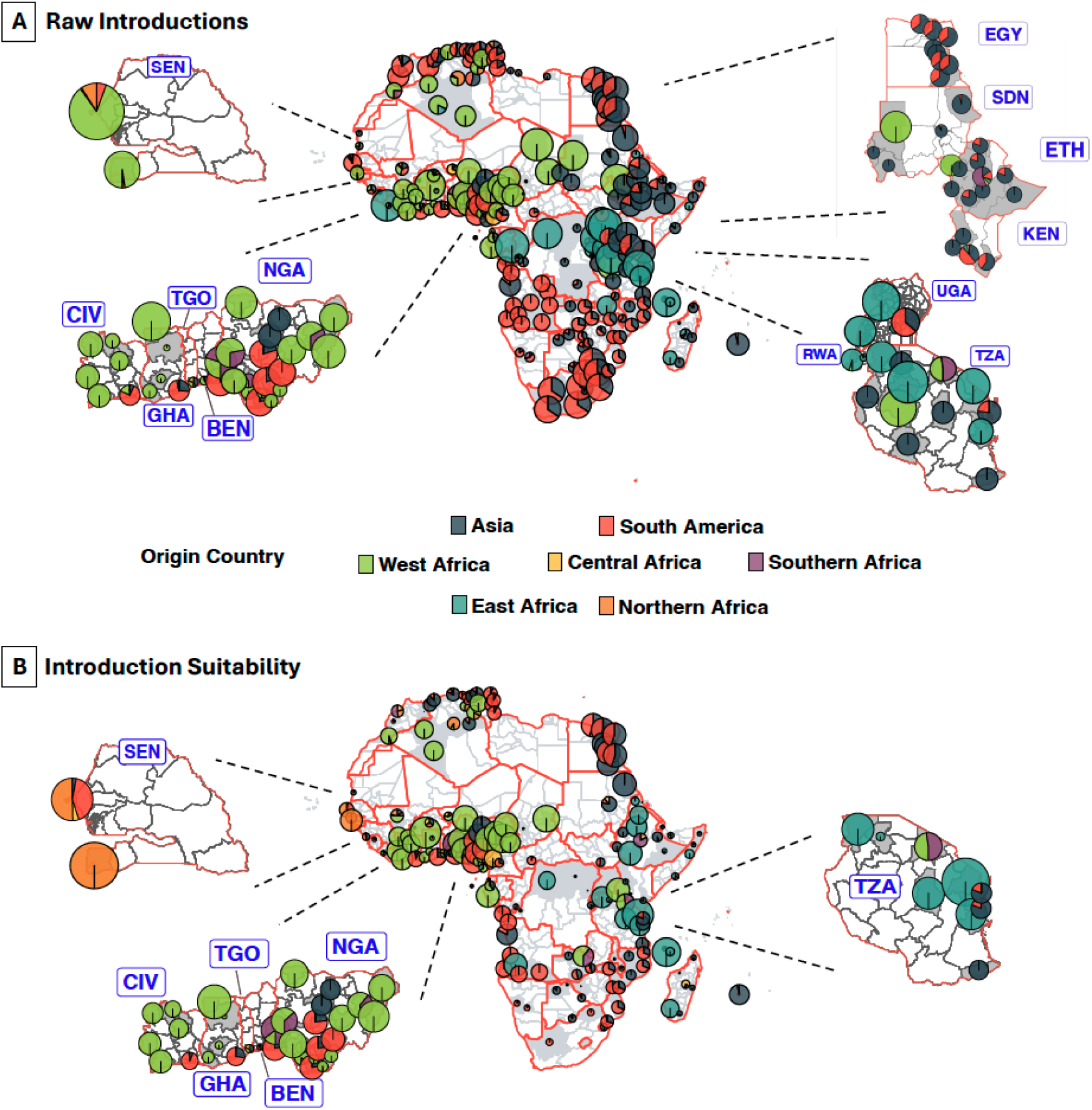
Overall proportional risk of importation into African countries before and after adjusting for period of persistence suitability (transmission suitability greater than 1). A) The proportion of risk from Asia,South America and African regions throughout the year for each district. The size of the pie charts are proportional to mean risk of importation across the year and the coloured pies represent the origin continent/region. We zoomed in on specific countries within the plot, namely: Ghana, Togo, Benin, Nigeria, Rwanda, Uganda, Senegal Tanzania, Egypt, Sudan, Ethiopia, and Kenya. B) The proportion of risk from Asia,South America and African regions across the year, for months with high persistence suitability. We zoomed in on specific countries within the plot, namely: Ghana, Togo, Benin, Nigeria, Senegal and Tanzania.

In our analysis, a noteworthy shift in risk proportionality emerged when focusing on introduction suitability compared to raw risk. For example, in the case of Dakar in Senegal, initially, 87.7% of the total risk was attributed to West African region, followed by 3.9% from South America and 6% from Northern Africa. However, when looking at periods of persistence suitability for Senegal, the risk distribution shifted and the proportion of risk attributed to Northern African regions, rose to 50.4%, followed by South America with 41.5%. This outcome underlines the importance of understanding not only the geographic origin of importation risks but also the specific temporal and spatial context in which the potential introductions can occur. Focusing on introduction suitability risk, countries in the West and East African regions exhibited significant risks of dengue importation. Notably, the highest risks for these countries were associated with regions within Africa itself (Figure 5).

**Figure 5:**
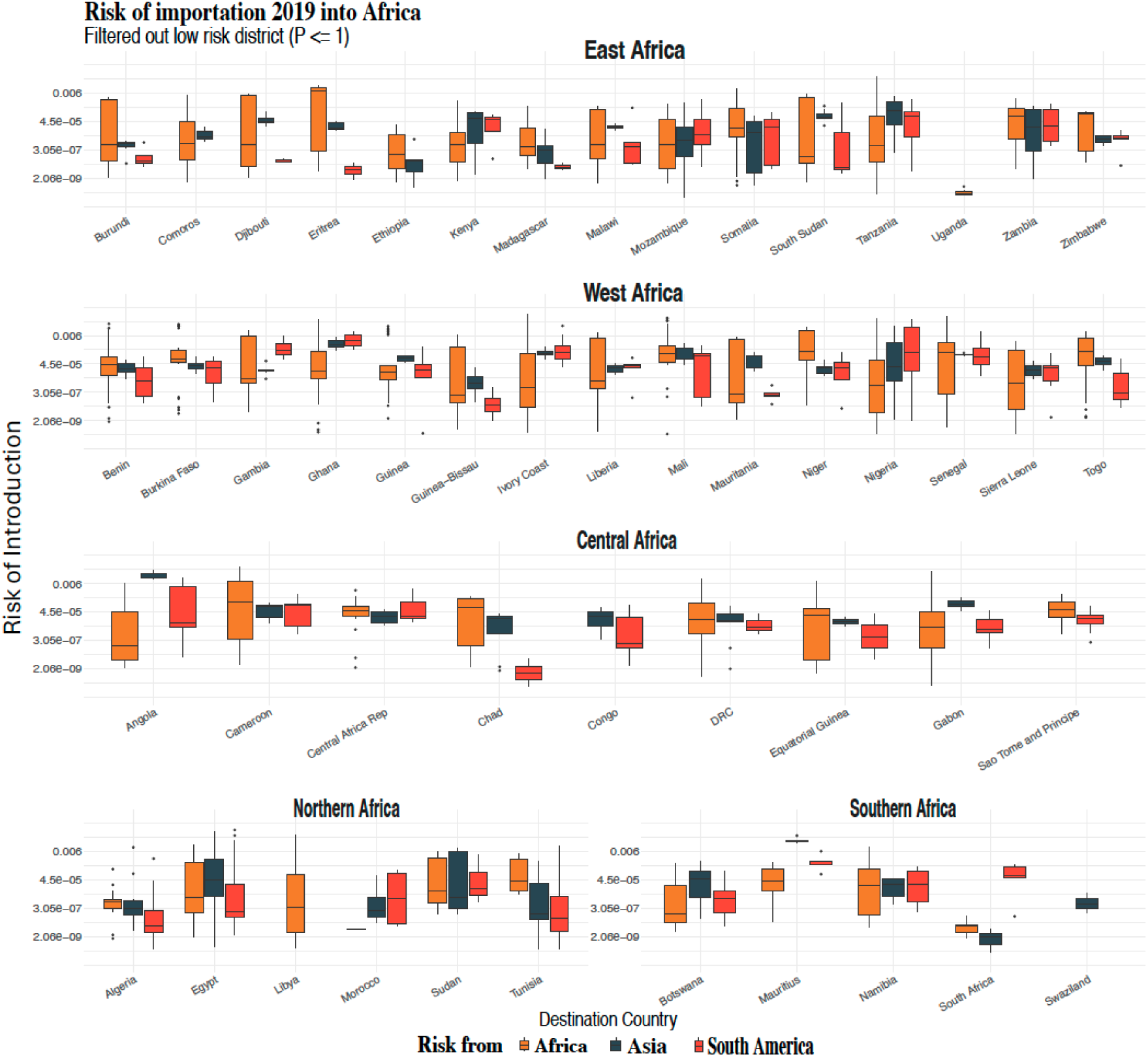
Risk of Dengue importation into various African countries, grouped by regions: East Africa, West Africa, Central Africa, Northern Africa and Southern Africa. Boxplot of risk across months and districts with high persistence suitability. Africa in orange, Aisa in blue and South America in red.

## Discussion

In this study, we investigated the temporal and spatial intersections between high risk of introduction and heightened local transmission suitability of dengue in African countries. The exploration of dengue introduction into Africa is imperative due to the potential implications for onward transmission. To this end, we constructed an importation suitability map to depict the spatial and temporal hotspots of dengue introduction and establishment into Africa from high incidence countries in Asia,Latin America, and regionally from other African countries.

We identified that the heterogeneity of the risk across Africa is dependent on the distribution of dengue activity across different provinces or states in the countries of high incidence and their connection to destination airports in Africa (see supplementary figure S8, p.9). Further, we found that seasonal variation plays a crucial role. For example, in India, major outbreaks tend to occur after the monsoon season and thus have a higher transmission suitability during these seasons (monsoon season spans from June to September) (33–35) and thus we also estimate a higher risk of dengue importation from India to African countries following the monsoon season (see supplementary figure S8, p.9).

This research presents an alternative risk assessment by considering modelled dengue transmission suitability (7) rather than reported incidence, compared to other studies using case incidence which might suffer from substantial underreporting (36–39). The correlation analyses revealed a robust association between transmission suitability and dengue cases for countries where high resolution spatiotemporal case records were available, with a significant correlation observed at a one-month lag (see supplementary figure S1, p.1). Our findings are consistent with previous research, demonstrating that 96% of municipalities in Brazil and 95% of provinces in Thailand exhibited incidence dynamics with 0-3 months time delays relative to estimated transmission suitability (7). Our correlation further supports the utility of using transmission suitability as a proxy for local dengue circulation.

Increased international travel and trade have facilitated the global spread of dengue from endemic to non-endemic regions, by enabling the movement of infected individuals and mosquito vectors between regions (6–8,10). The risk flow results presented here highlight the substantial risks of dengue introduction into certain parts of the African continent from Asia, South America, or regionally from other African countries through human mobility. We show that the risk of importation to African countries is highly heterogeneous. The results produced by our model suggest that major airports in South Africa, Egypt, Morocco, Kenya, Seychelles and Mauritius, are potential hotspots for the importation of dengue-infected travellers from high-incidence countries. However, the analysis also revealed important geographical nuances: for example, the broader Eastern African region, and parts of Southern and Africa, are confronted with elevated risks of dengue importation from Asia, while Central, and segments of Northern and Southern Africa are more susceptible to introductions from South America (albeit often not suitable introductions). This is consistent with a recent phylogeographic study which found that all four dengue serotypes were introduced on multiple occasions to Africa, primarily from South or Southeast Asia since the 1940s (40). This highlights the significance of our study in obtaining high-resolution and recent temporal and spatial trends for the vulnerability of Africa to dengue introductions.

In contrast, our analysis of the risk of importing dengue from within the continent reveal areas of the continent where a substantial portion of the risks tends to come from within the region, underscoring the importance of intra-continental transmission dynamics. This is estimated to be the major driver of cross-country introduction in West Africa, and to a lesser extent in East Africa. This is supported by recent phylogeographic reconstructions which showed frequent introductions and reintroductions of the dengue virus within the western African region, as well as similar patterns in the eastern part of the continent (40,41). While Asia represents the second largest contributor to dengue risk, particularly impacting the East, West and Central African regions, the internal risk within Africa remains a primary concern. To put this in context, Africa exhibits relatively similar or closely aligned spatial characteristics in transmission suitability across its regions and therefore, it is expected that intra-continental risk remains predominant. This spatial similarity reinforces the interconnectedness of African regions in the context of disease transmission dynamics. On the other hand, there is also a substantial contribution from Asia particularly in East Africa, and towards the island nation of Mauritius, indicating that these countries are vulnerable to introductions both from within the continent and from Asia.

It remains crucial not to disregard regions with high population density but that currently express low transmission suitability, such as Ethiopian highlands or high altitude regions of Tanzania. Despite their dense population, these regions have historically shown low transmission suitability values. However, with the potential impacts of future global climate change, these areas may become increasingly susceptible to local dengue outbreaks. By integrating appropriate travel models, this approach can be adapted to predict dengue risk under various climatic conditions, providing valuable insights for future public health planning.

To translate the high risks of introduction into the notion of introduction suitability, that is considering potential introductions that are highly likely to cause onward transmission in the destination countries, it was important to understand the specific temporal and spatial patterns of the risk flow estimates. The temporal analysis of estimated importation risks and local transmission suitability revealed the optimal times to conduct increased surveillance, as the times when the risks of introduction and the potential for transmission are simultaneously high. Therefore, the temporal synchrony between estimated introduction risks and transmission suitability has important implications for which viral introductions are predicted to have the ability to contribute to local outbreak risks.

Epidemiological and genomic surveillance are critical public health measures to mitigate disease burden, and also to understand the spread and evolution of pathogens. At present, there is no approved antiviral treatment for dengue and supportive care is the only option (17–19). However, two dengue vaccines have shown great promise (Takeda and Butantan vaccines) (20). Genomic surveillance can provide critical information about viral transmission landscape prior to upcoming dengue vaccine rollouts. To determine the optimal locations for conducting genomic surveillance in Africa, it is also important to have a quantitative understanding of the actual risk of a dengue virus epidemic at country level, particularly given limited resources to implement surveillance programs. This study’s findings point to the fact that emphasis should be placed on targeted monitoring during periods when the likelihood of risks escalating into an outbreak is higher, and within areas prone to introductions during time of persistence suitability. In turn, in light of introduction risks, genomic surveillance for dengue enables public health officials to monitor the circulating serotypes while remaining vigilant for the emergence of novel strains or genotypes. In the particular case of dengue, introductions could lead to outbreaks with worsened disease outcomes in a population that has already been exposed to a different serotype of the virus, and depending on the temporal sequence of serotype circulation. This is a growing concern in Africa and elsewhere given the number of countries in which dengue is now well established and serotype co-circulation is becoming universal. Importantly, our modelling approach can also be used for other continents, namely Europe and North America where dengue epidemic activity is increasing.

This study should be interpreted in light of several limitations. First, this study presents relative introduction and establishment risks calculated from ecological, statistical, and mathematical models but does not present direct estimates of the number of expected introductions. Further, due to paucity of genomic and traveller testing data the models presented here have yet to be validated. In countries that perform disease surveillance of incoming travellers, the history of travel of positive dengue cases could serve as validation data to this type of modelling study. Rapidly expanding volume of available dengue genomes will soon make it possible to reconstruct high resolution transmission dynamics to validate introduction models presented here. Additionally, the measure of transmission suitability used for the source locations does not account for specific control measures in place, potentially impacting the accuracy of the estimated dengue incidence. It also does not take into account the presence and abundance of mosquito vectors. Despite this limitation, the chosen transmission suitability index P seems to produce realistic estimates for dengue incidence in source locations, considering variations in testing and reporting practices globally. Thirdly, while our findings are founded upon the current travel networks, it is imperative to acknowledge the dynamic nature of travel patterns. Given the expansive reach of air travel and the transformative impact observed during the COVID-19 pandemic, we must remain vigilant to the possibility of shifts in dengue introduction risks. This is particularly relevant as more areas are becoming suitable for transmission of dengue. Furthermore, the connectivity between African countries is only captured here through air travel data, while the true connection between neighbouring countries would also be dependent on road travel networks and information about the porosity of borders, data which is much more difficult to obtain. Finally, models like the one employed in this study rely on air travel data and do not account for the role of trade in moving infected mosquitoes between countries (e.g. in tyres); this should be considered in future work.

In conclusion, this study provides valuable insights into the complex dynamics of dengue importation risks into Africa, while making a distinction as to the source of those potential introductions. The incorporation of estimated transmission suitability for dengue and population density in risk assessment enhances the accuracy of predictions. The temporal and spatial analyses highlight specific regions and times that warrant intensified surveillance and public health interventions given the likelihood of potential introductions that would lead to local outbreaks. These findings contribute to a more nuanced understanding of global dengue dynamics, and importantly focus on informing further surveillance on the Africa continent and globally.

## Data Availability Statement

Proprietary air travel data are commercially available from the International Air Transport Association (https://www.iata.org/) databases. Transmission suitability (index P) estimates are available at https://doi.org/10.6084/m9.figshare.21502614. Risk estimates computed from this study can be found at:

https://github.com/CERI-KRISP/Dengue_Importation_Risk_Modelling.git

## Contributors

H.T. and J.P. conceptualised and designed the study. J.P. analysed data, executed all primary data visualizations, and wrote the original draft. V.C. accessed the travel data. Y.R. collected and curated epidemiological data. M.D., T.d.O, V.C. and M.U.G.K. interpreted data. J.L. and J.L.-H.T. contributed to data analysis and visualisation. C.B. and T.d.O acquired funding for this project. H.T., M.D., T.d.O supervised the study. All authors had full access to all the data in this study. All authors reviewed and edited the final draft. All authors had final responsibility for the decision to submit for publication.

## Acknowledgements

CERI and KRISP are supported in part by grants from the Rockefeller Foundation (HTH 017), the National Institute of Health USA (U01 AI151698) for the United World Antiviral Research Network (UWARN), and the INFORM Africa project through IHVN (U54 TW012041), Global Health EDCTP3 Joint Undertaking and its members as well as Bill & Melinda Gates Foundation (101103171), European Union’s Horizon Europe Research and Innovation Programme (101046041), the Health Emergency Preparedness and Response Umbrella Program (HEPR Program), managed by the World Bank Group (TF0B8412), the Medical Research Foundation (MRF-RG-ICCH-2022-100069), and the Wellcome Trust (228186/Z/23/Z). M.U.G.K. acknowledges funding from The Rockefeller Foundation (PC-2022-POP-005), Google.org, the Oxford Martin School Programmes in Pandemic Genomics & Digital Pandemic Preparedness, European Union’s Horizon Europe programme projects MOOD (#874850, also V.C.) and E4Warning (#101086640), a Branco Weiss Fellowship and Wellcome Trust grants 225288/Z/22/Z, 226052/Z/22/Z & 228186/Z/23/Z, United Kingdom Research and Innovation (#APP8583) and the Medical Research Foundation (MRF-RG-ICCH-2022-100069).. J.L.-H.T. is supported by a Yeotown Scholarship from New College, University of Oxford. V.C. acknowledges funding from Horizon Europe grants ESCAPE (101095619) and VERDI (101045989); EU Horizon 2020 grant MOOD (H2020-874850, paper 874850, also to M.U.G.K.).The content and findings reported herein are the sole deduction, view and responsibility of the researcher/s and do not necessarily reflect the official position and sentiments of the funding agencies.

## Supplementary Appendix

**Figure S1:**
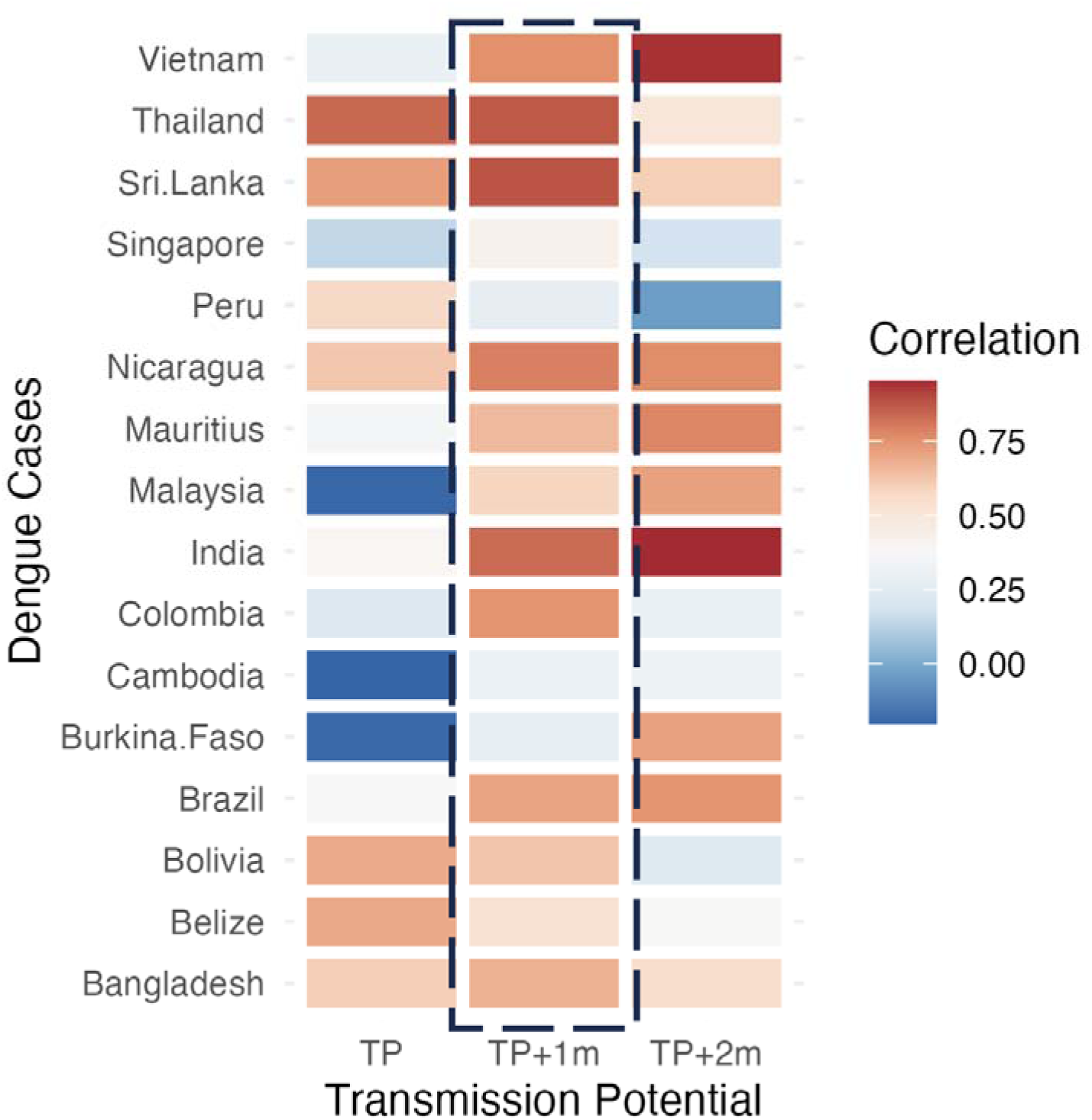
Temporal correlation between monthly dengue cases from different origin countries to Transmission potential without and with 1 and 2 months lag (p-values ranging from 0.001 to 0.65).

**Figure S2:**
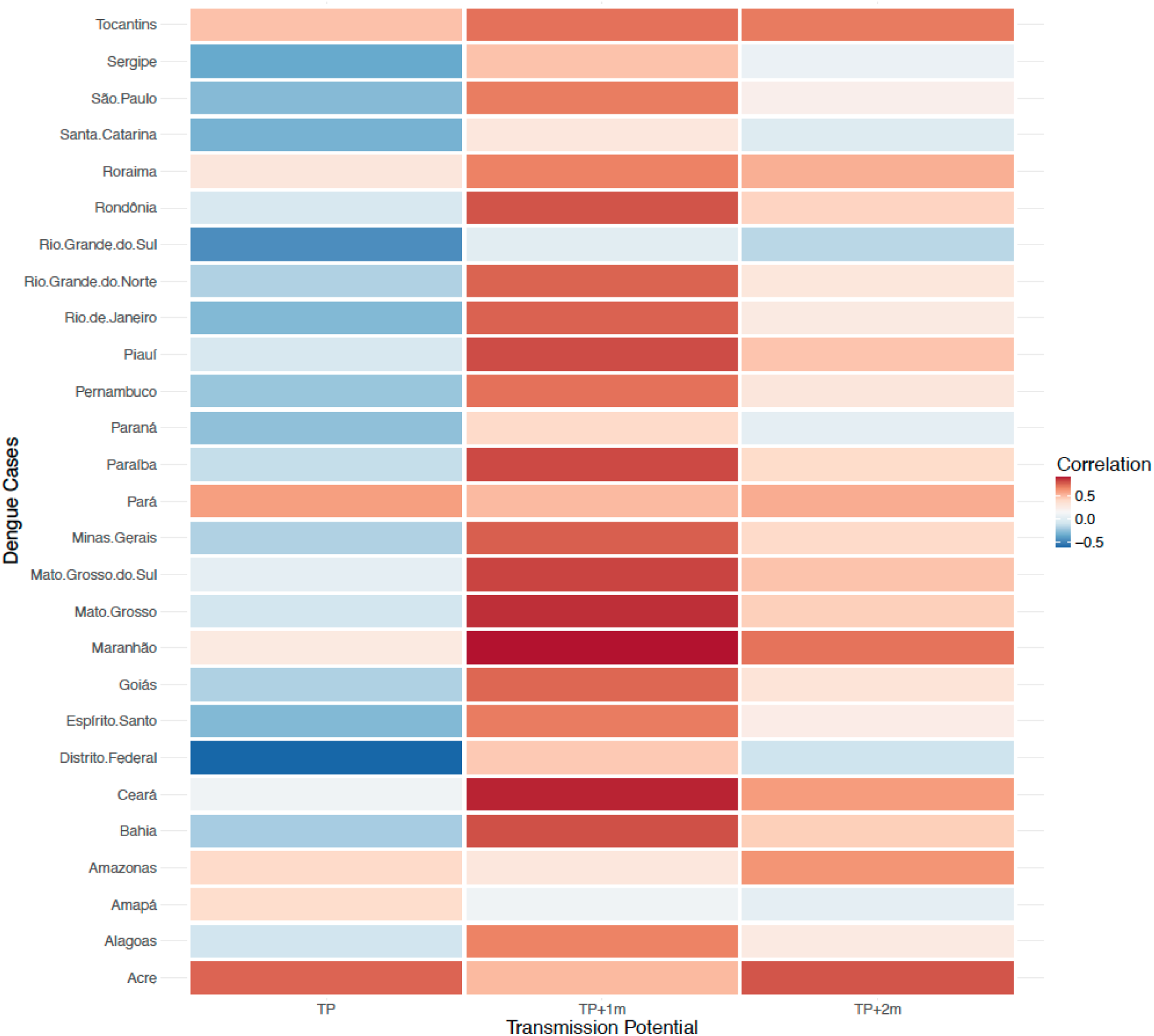
Temporal correlation between monthly dengue cases from different states in Brazil to Transmission suitability without and with 1 and 2 months lag. The p-values for the correlations ranged from 0.0000103 to 0.671.

**Figure S3:**
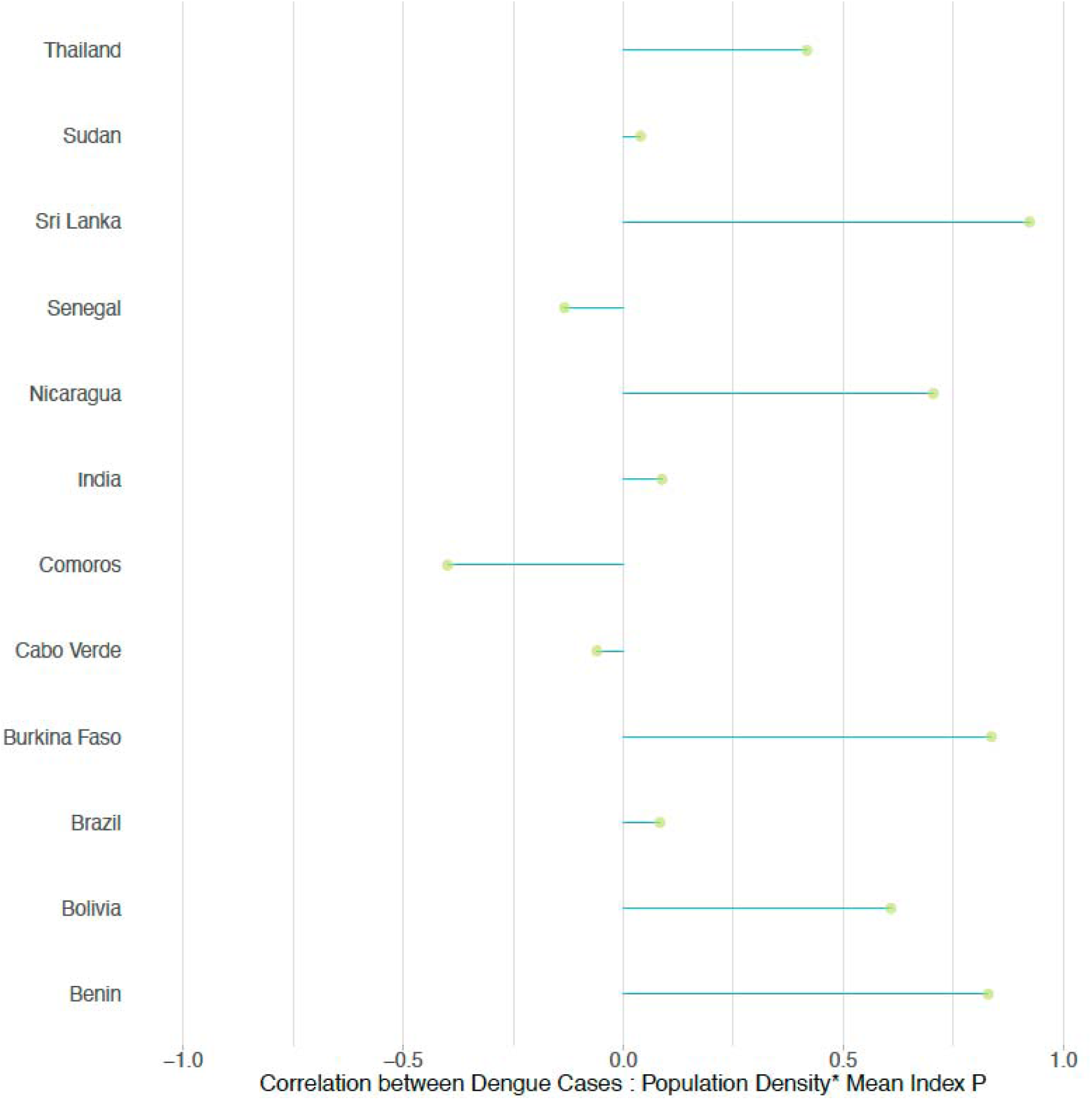
Correlation between Dengue Cases and Population Density multiplied by Mean Index P: This plot illustrates the relationship between the number of dengue cases and the interaction of population density with the mean transmission potential (index P).

**Figure S4:**
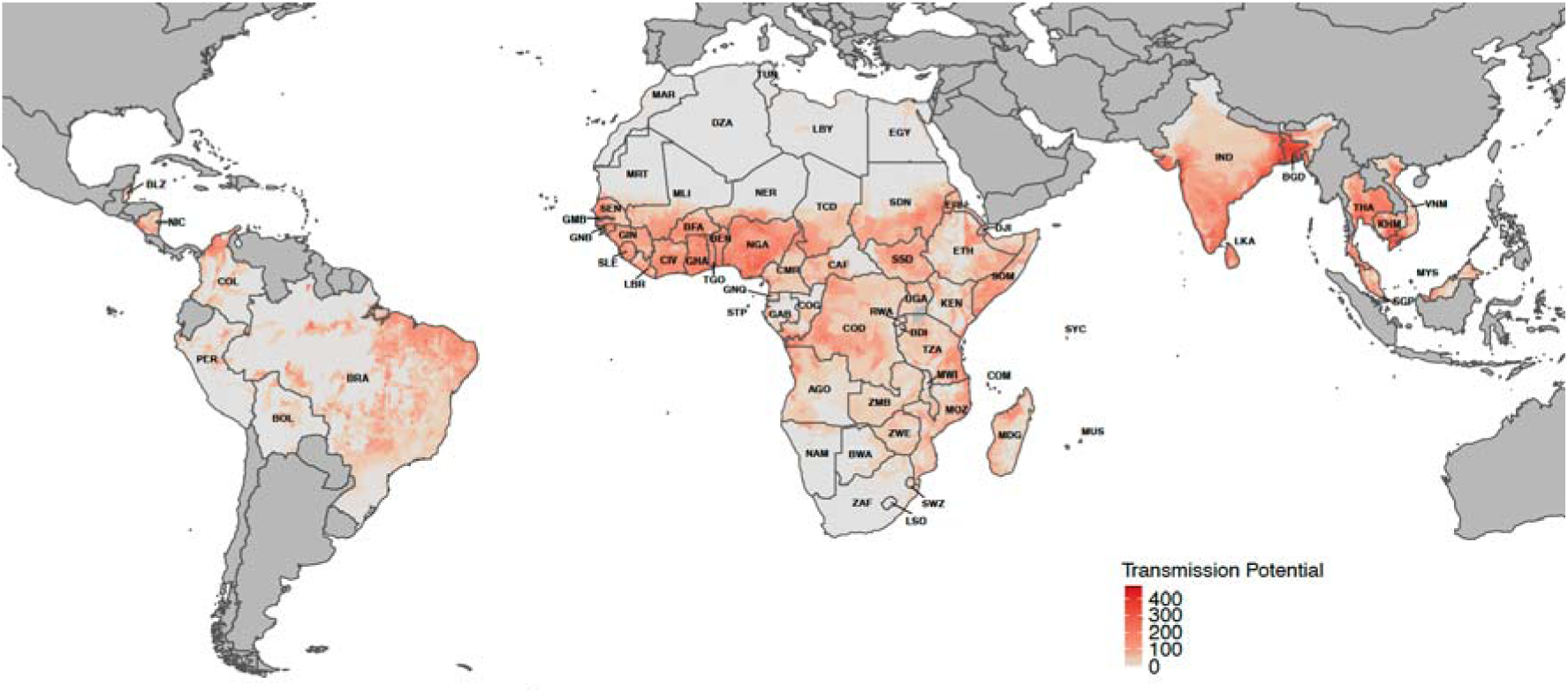
The figure shows the composite index of transmission suitability multiplied by Population Density (t_i), used in this study. Here we only show for the 14 countries of high incidence and the African continent, i.e., only for the origin and destination countries used in this study.

**Figure S5:**
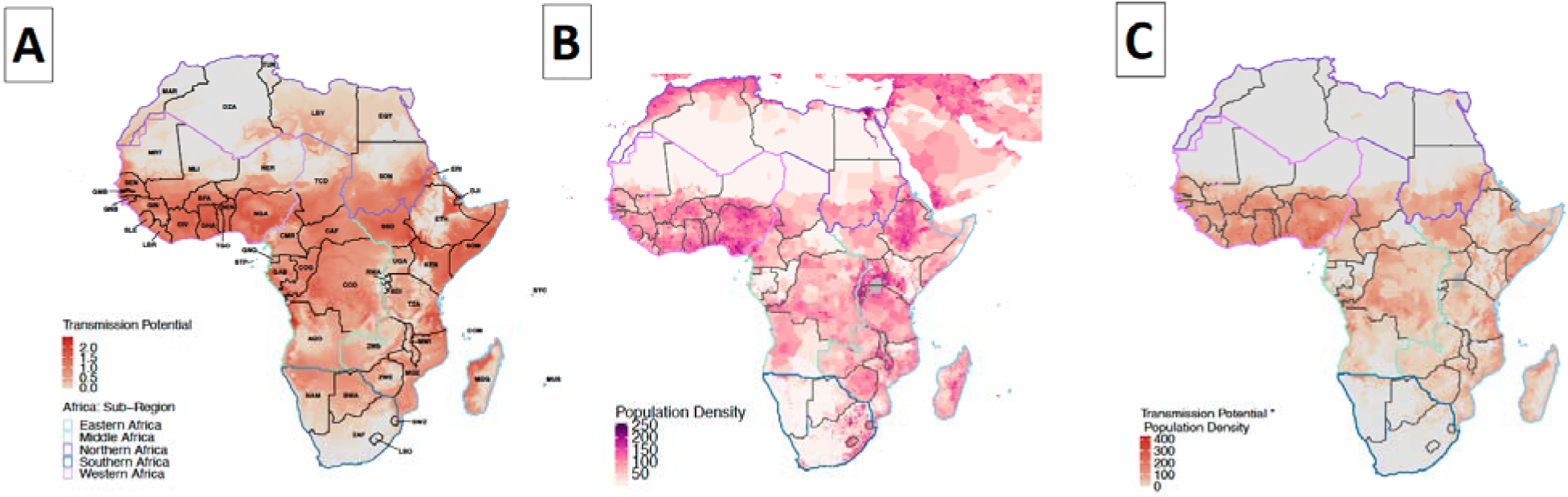
A) The map background uniformly represents the transmission suitability of dengue (mean index P) across Africa, with the continent divided into five distinct regions. B) The population density across Africa and C) Transmission suitability multiplied by population density for Africa. We also divide the African continent into 5 regions: Eastern (light blue), Middle (Green), Northern (Purple), Southern (dark blue) and Western Africa (pink) which are outlined on the map.

**Figure S6:**
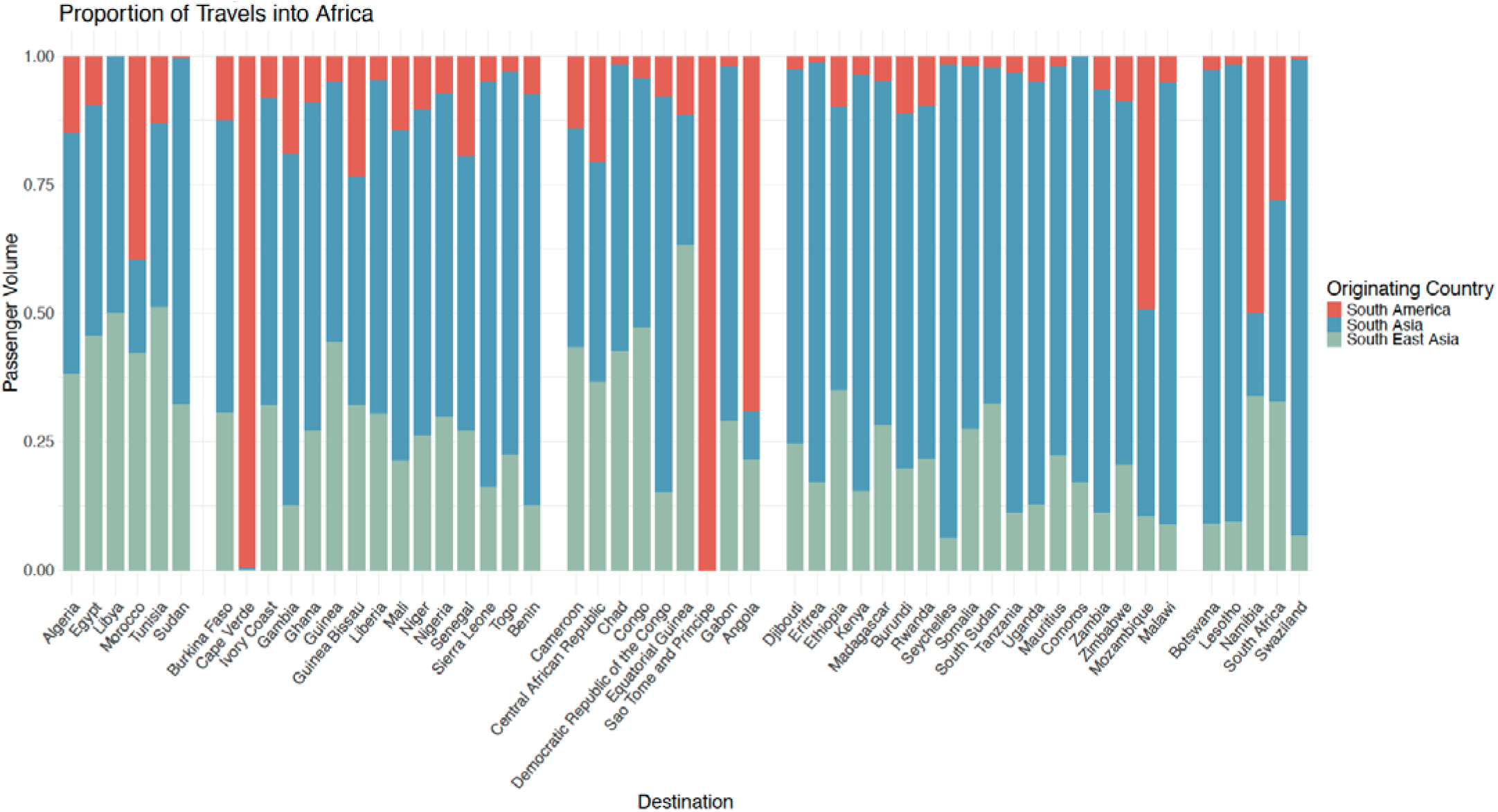
This figure depicts the proportion of travel volumes from Asia, Southeast Asia, and South America to African countries over the course of one year.

**Figure S7:**
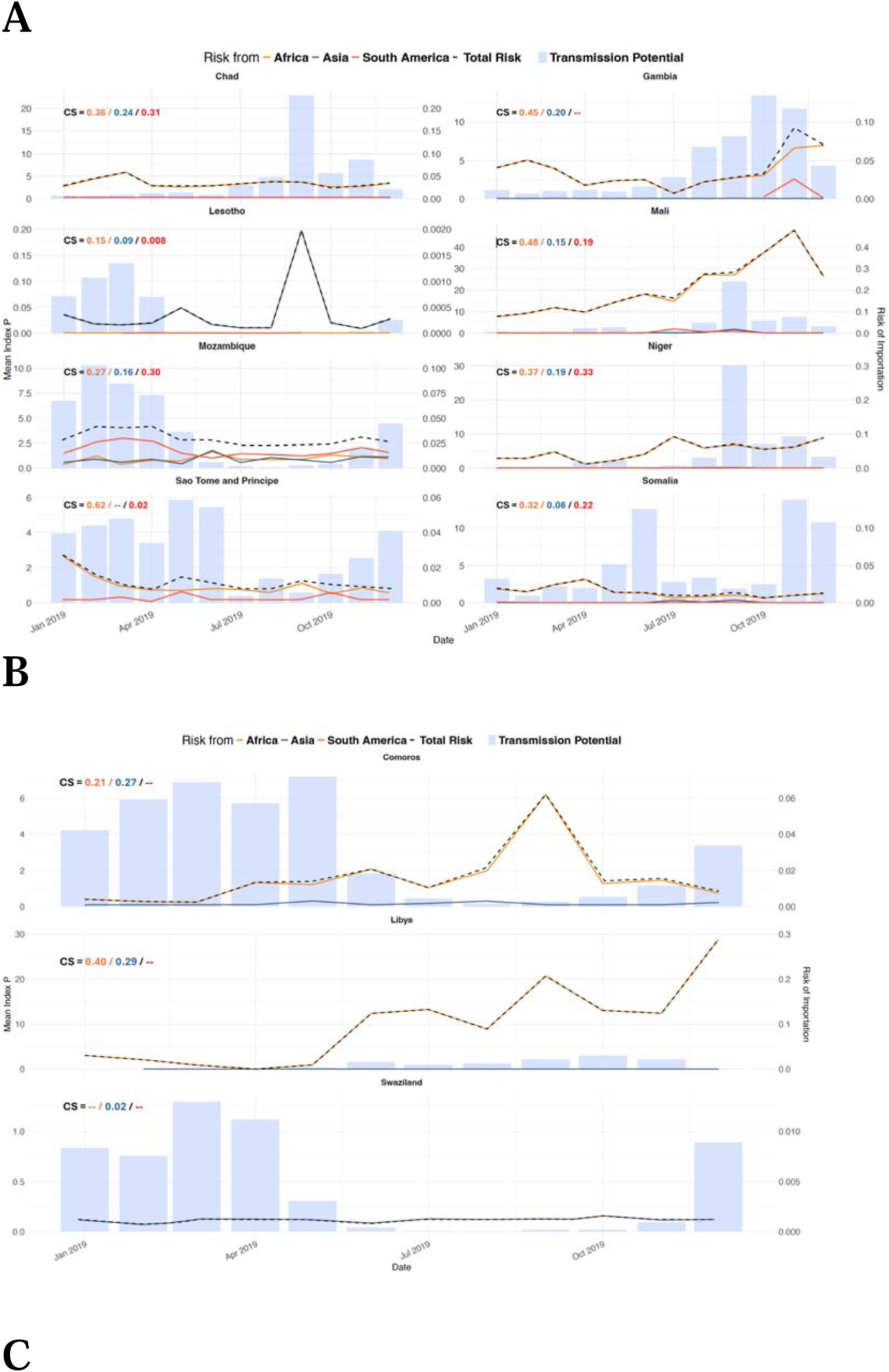
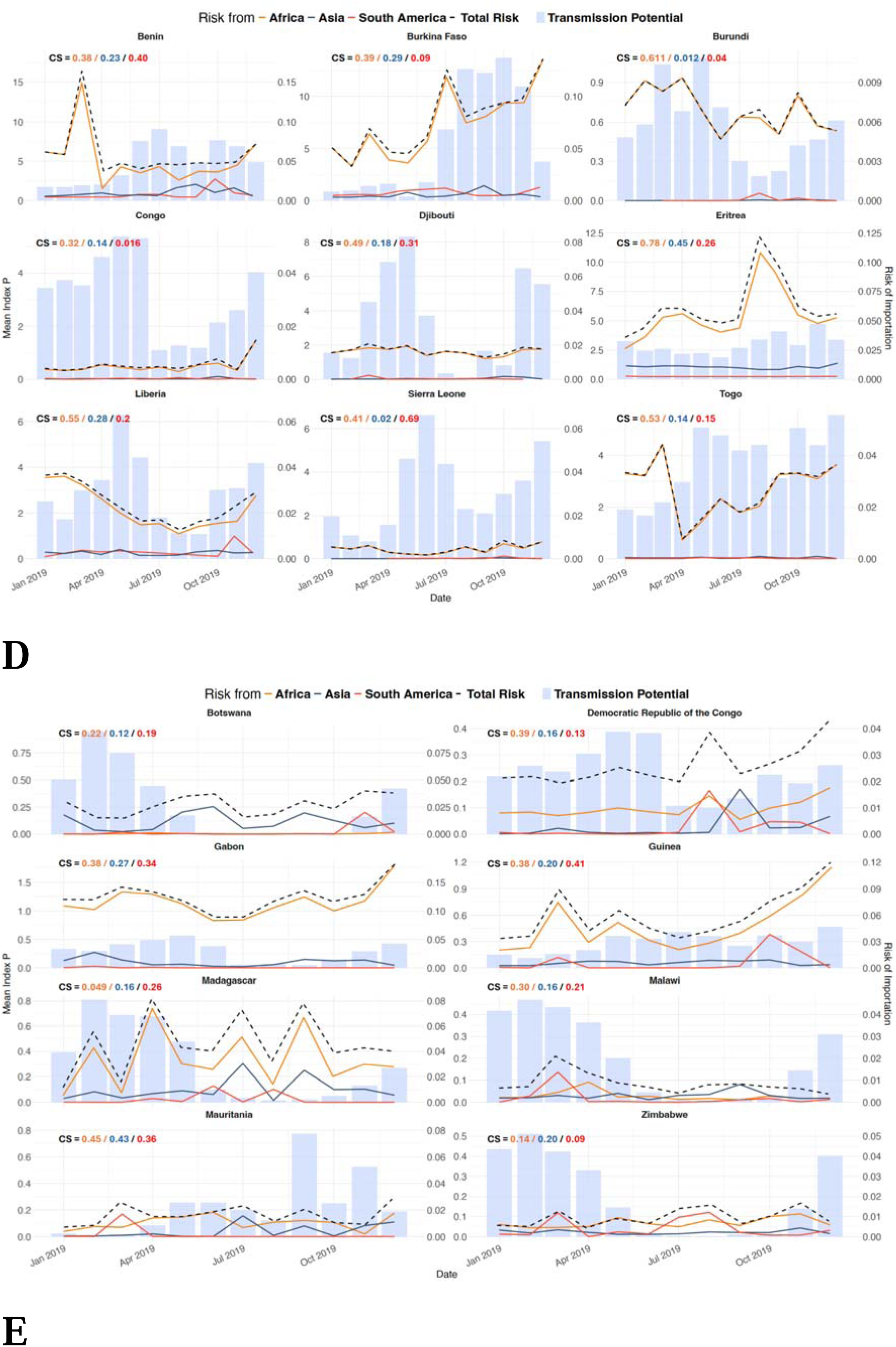
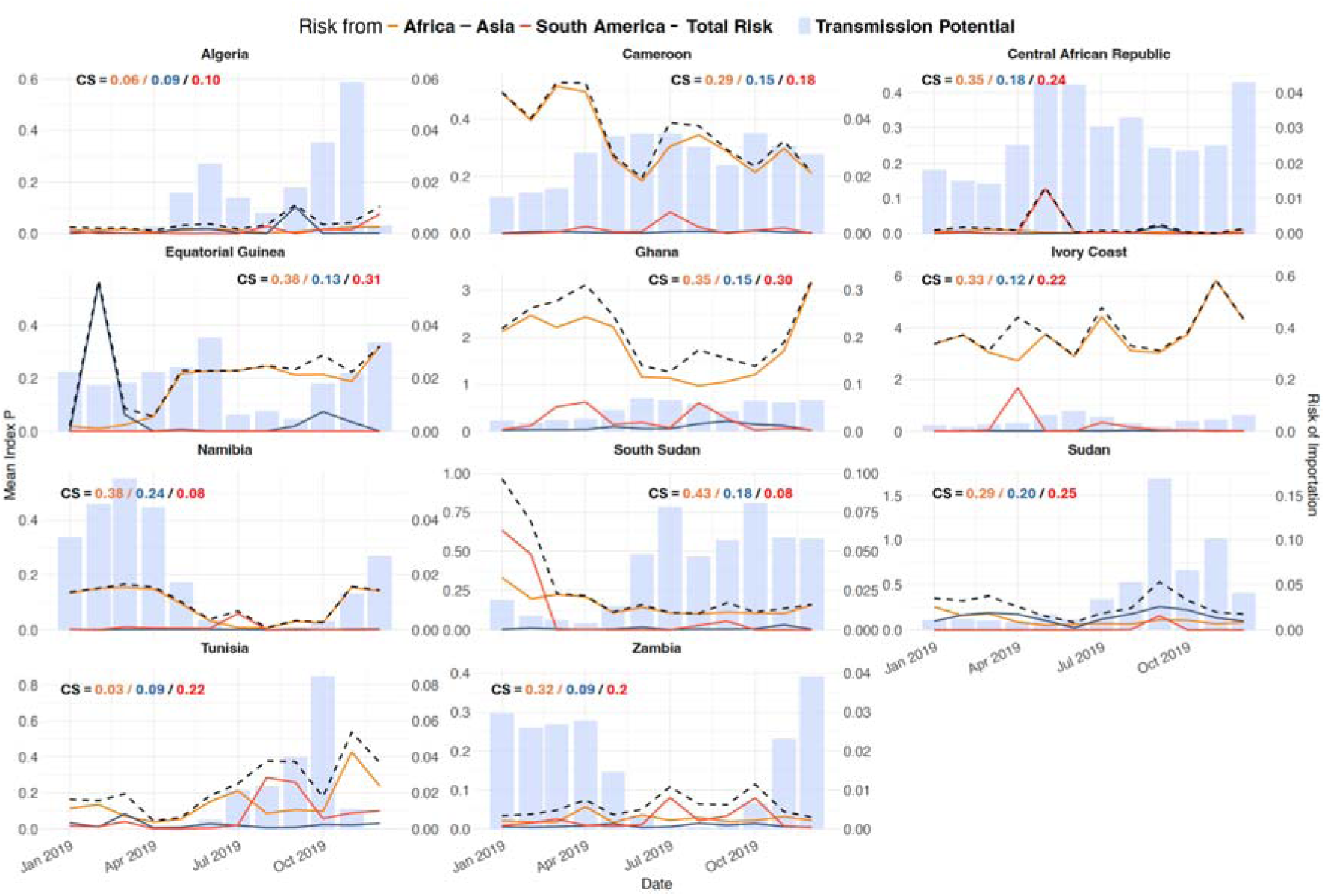
Time-varying risk of introduction into African countries in 2019 from Asia (blue line) and South America (red line) along with time-varying transmission suitability index nationally. The combined risk is represented by the black dotted line. Panel A) Chad, Gambia, Lesotho, Mali, Mozambique, Niger, Sao Tome and Principe, Somalia. B) Comoros, Libya, Swaziland. C) Benin, Burkina Faso, Burundi, Congo, Djibouti, Eritrea, Guinea Bissau, Liberia, Sierra Leone. D) Botswana, DRC, Gabon, Guinea, Madagascar, Malawi, Mauritania, Zimbabwe. E) Algeria, Cameroon, Central African Republic, EquatorialGuinea, Ghana, Ivory Coast, Namibia, South Sudan, Sudan and Tunisia.

**Figure S8:**
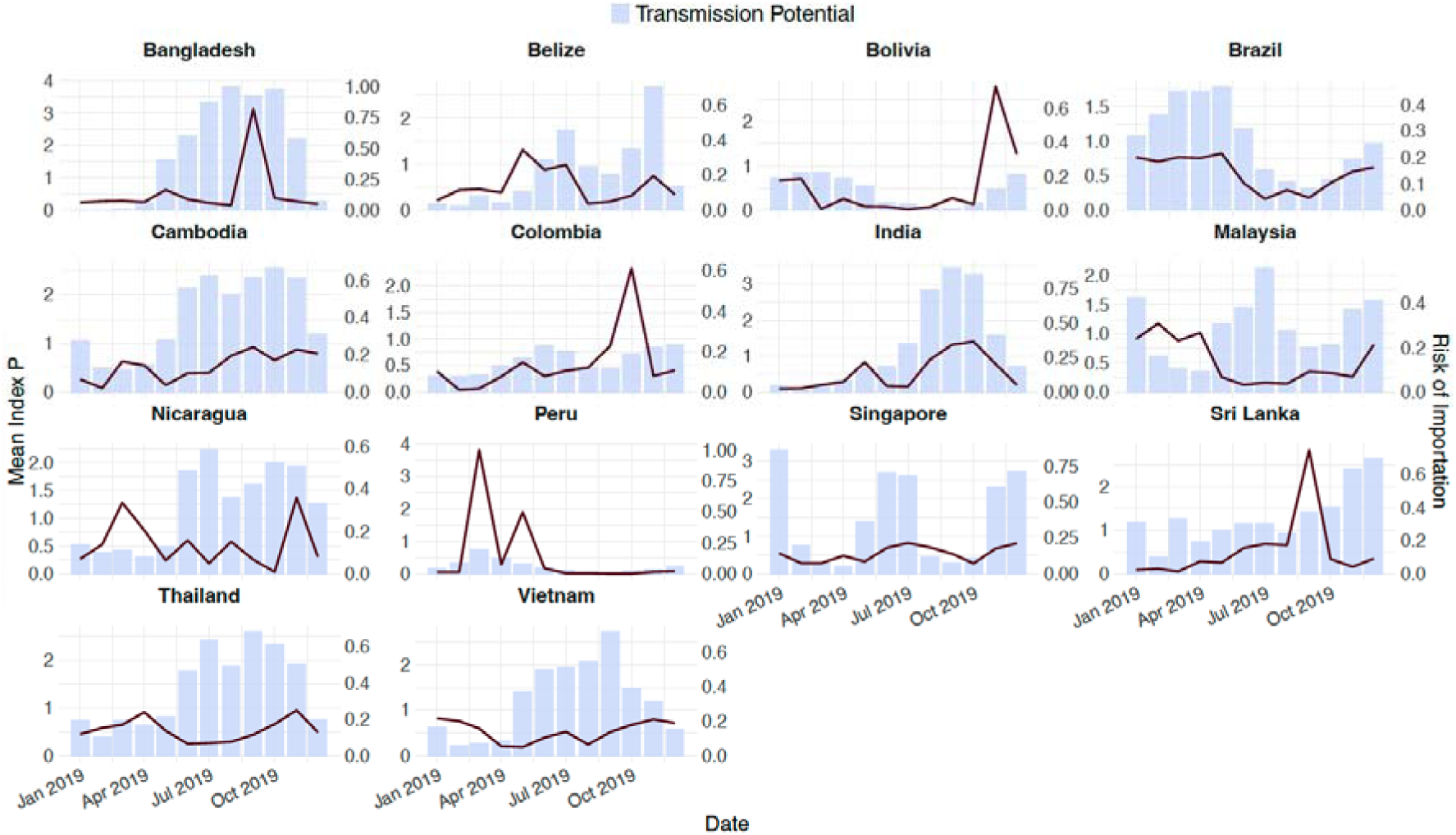
Risk of Exportation from countries of high incidence (n=14) considered in this study overlaid across their respective transmission suitability across the year.

**Table S1:**
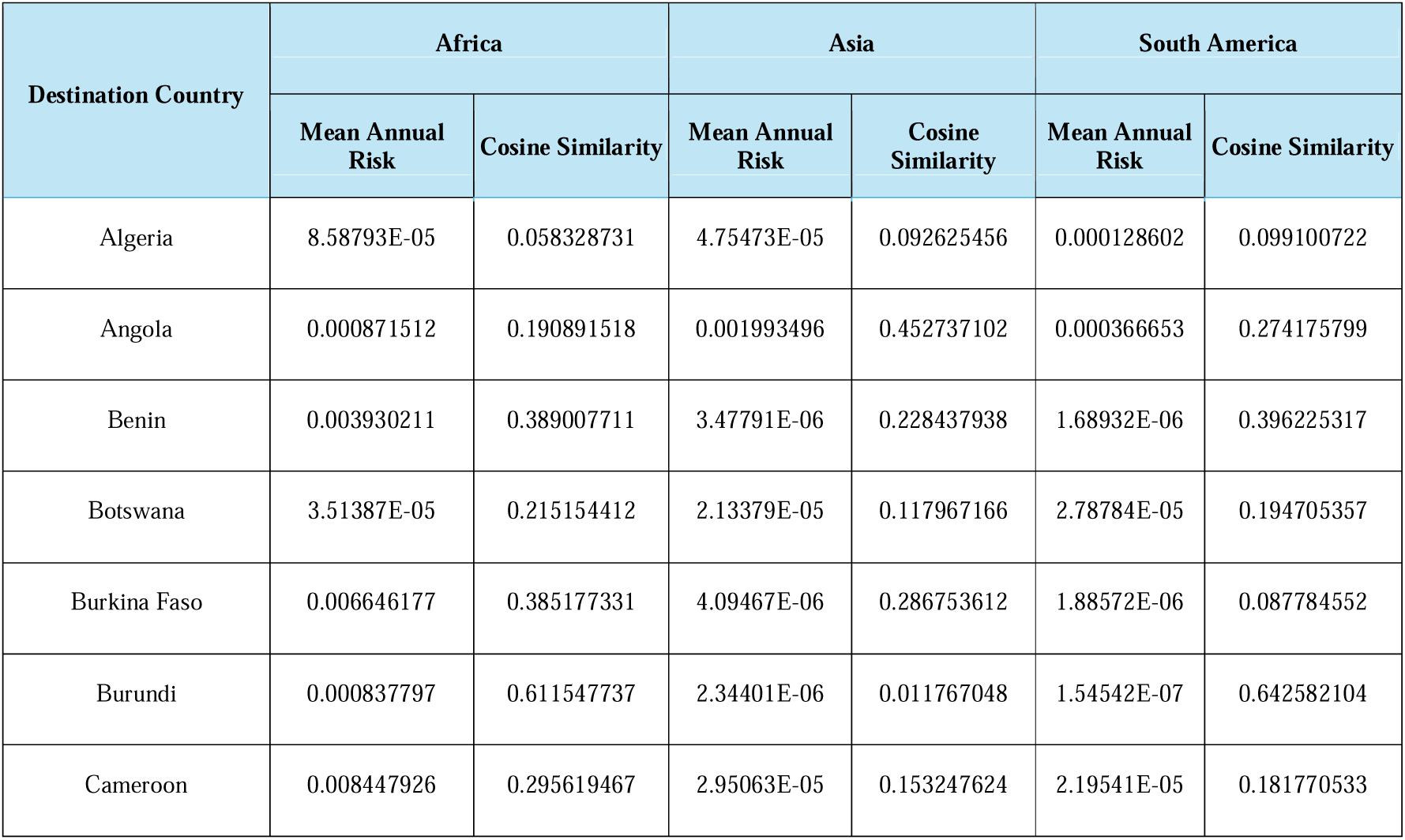
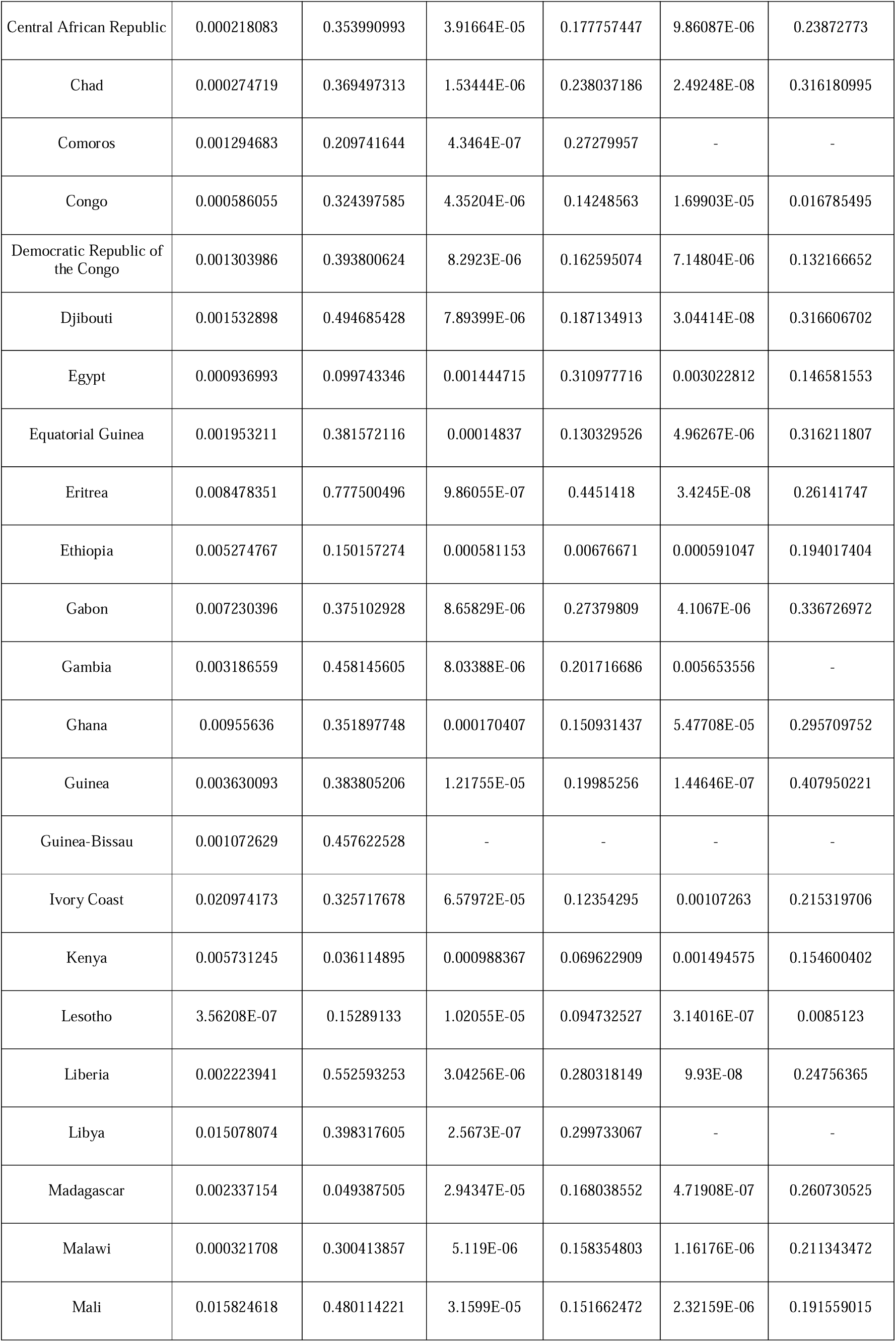
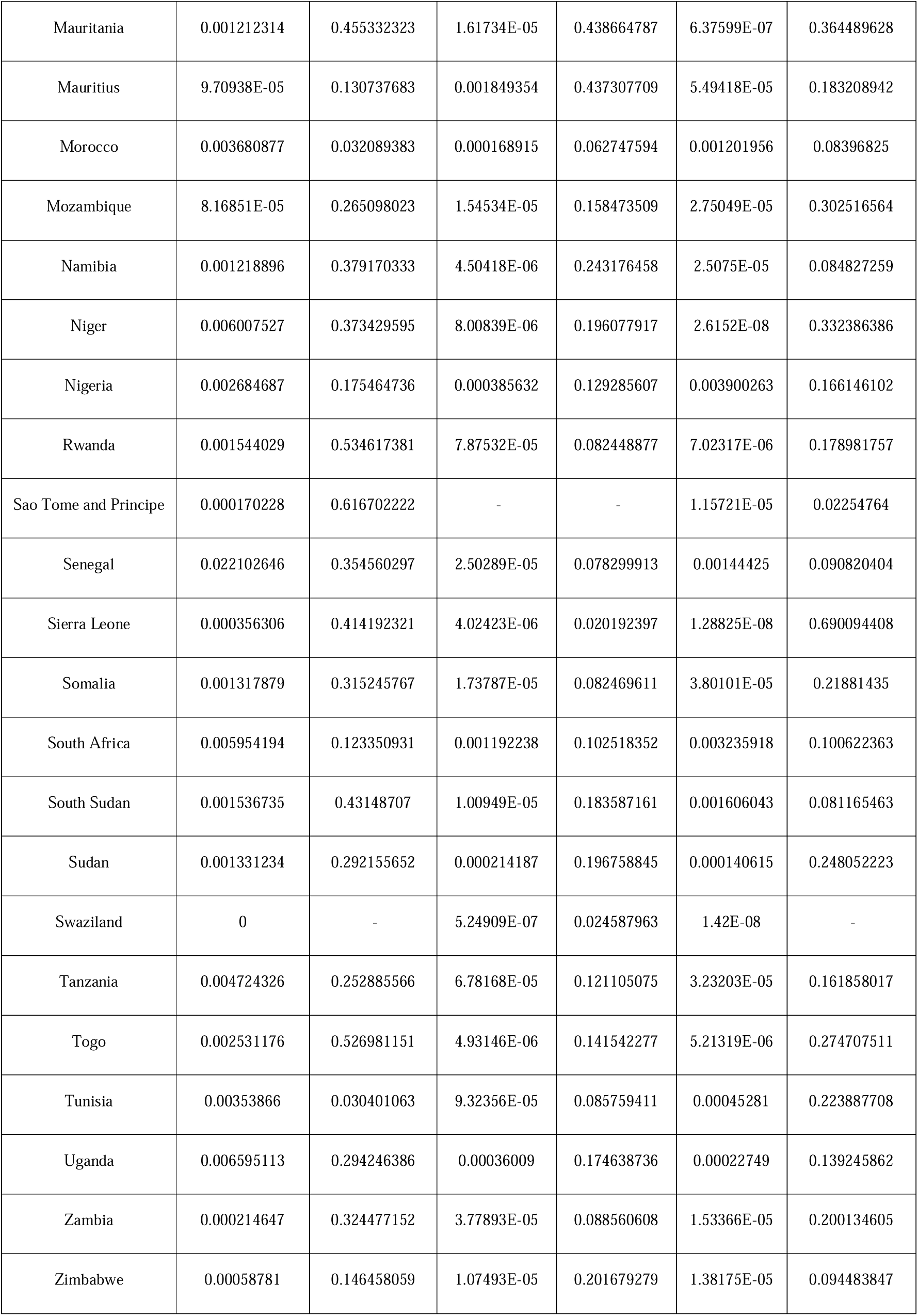
Annual risk of dengue importation into various countries and the cosine similarity between the risk of importation and the local transmission suitability. The cosine similarity values highlight the level of synchrony between the timing of importation risk and transmission suitability.

