## Supplementary Information for "Assessing Dengue Virus Importation Risks in Africa: A Climate and Travel-Based Model"

### Supplementary Appendix

#### Figure S1

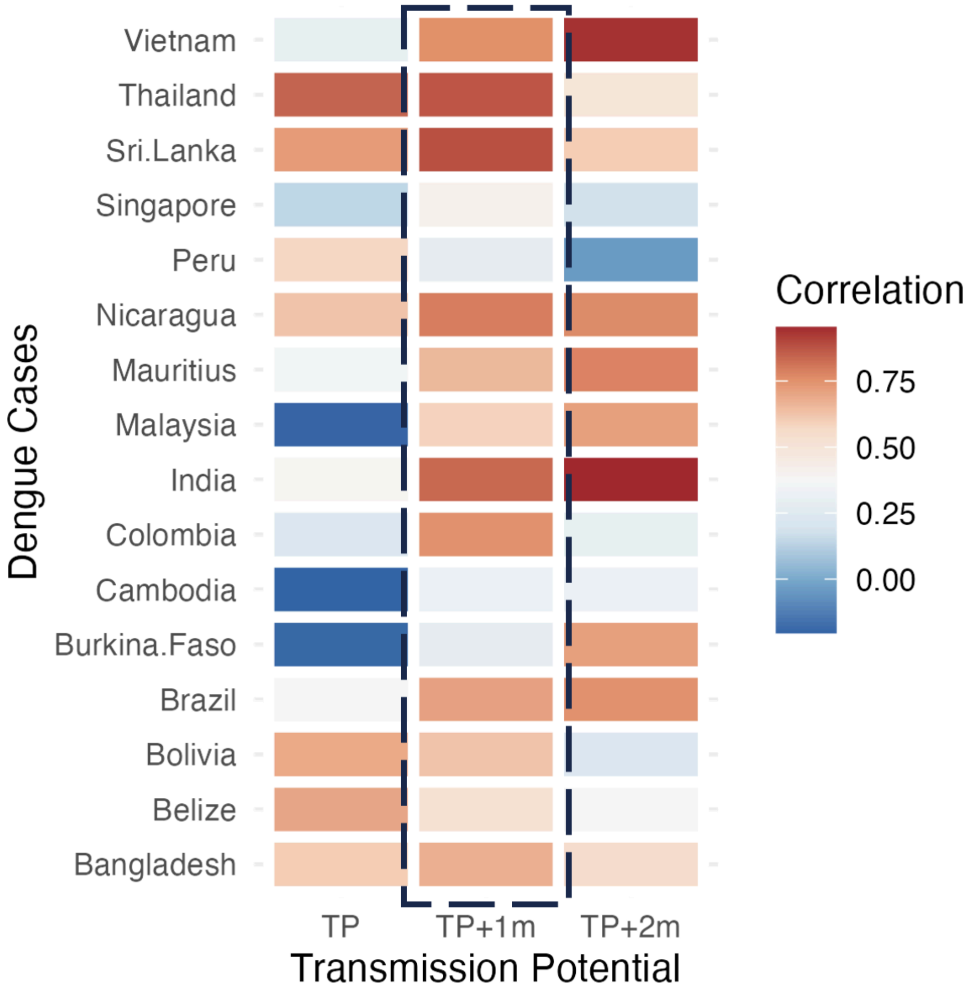


Figure S1: Temporal correlation between monthly dengue cases from different origin countries to Transmission potential without and with 1 and 2 months lag (p-values ranging from 0.001 to 0.65).

### Figure S2

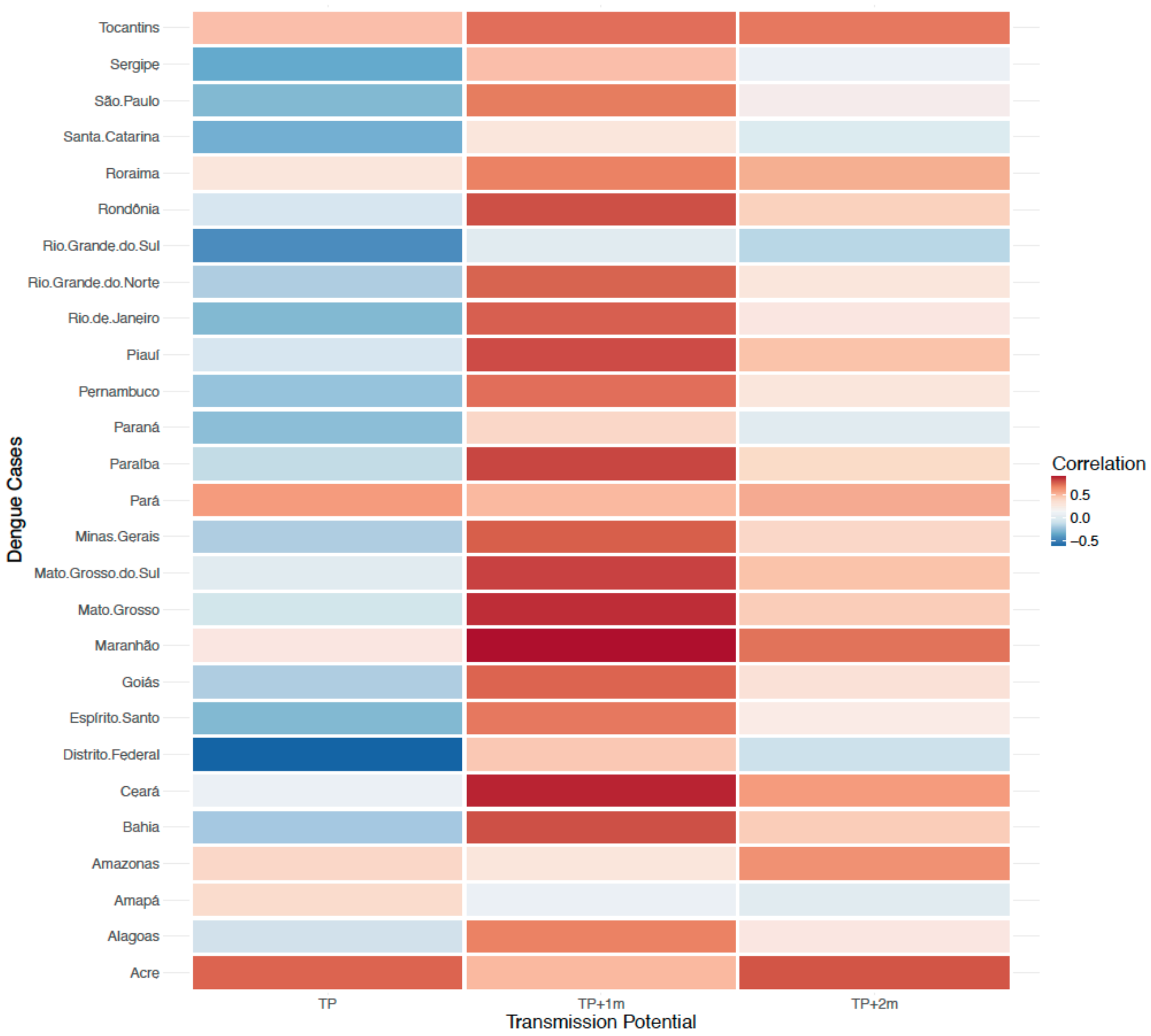


Figure S2: Temporal correlation between monthly dengue cases from different states in Brazil to Transmission suitability without and with 1 and 2 months lag. The p-values for the correlations ranged from 0.0000103 to 0.671.

#### Figure S3

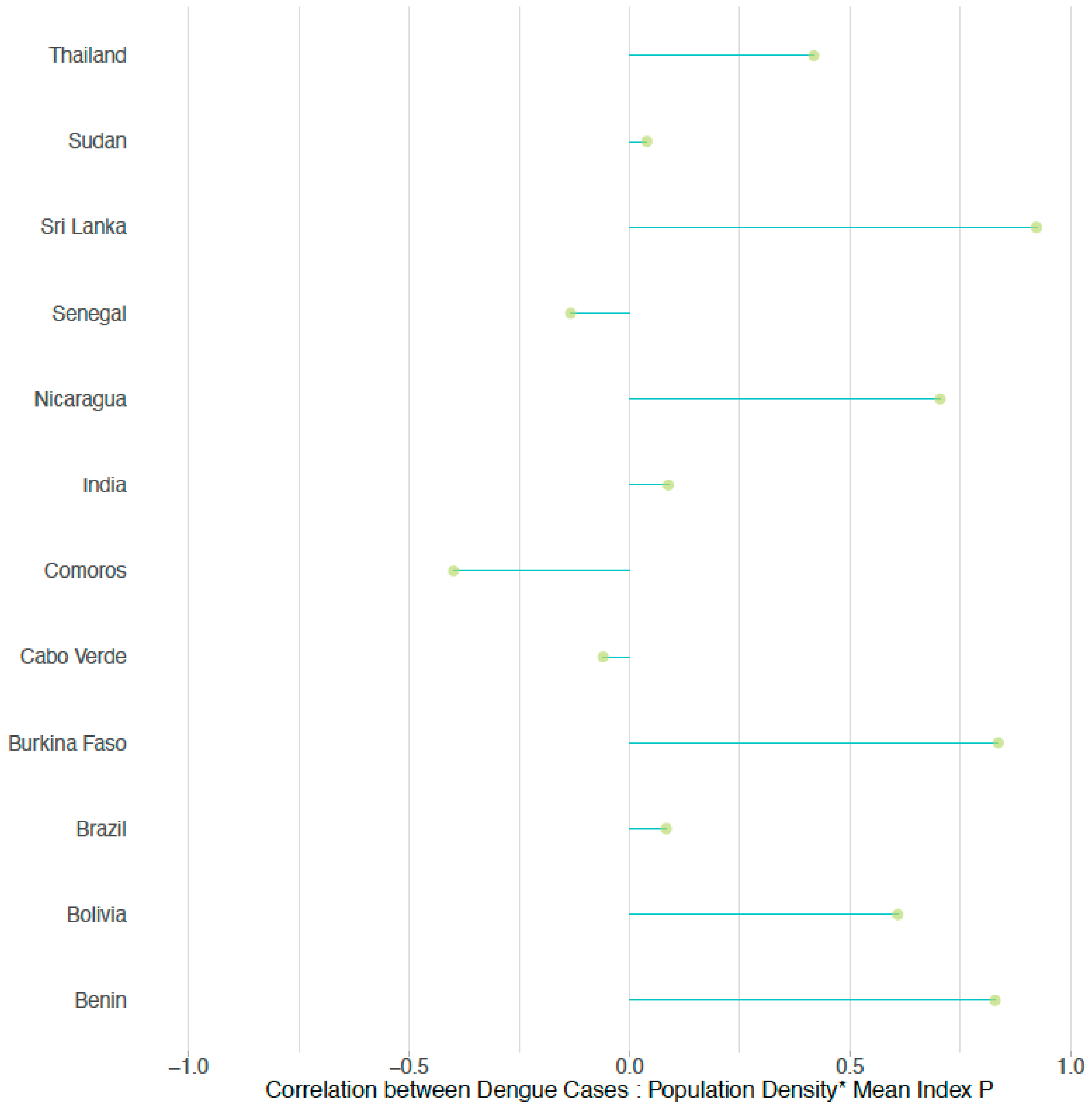


Figure S3: Correlation between Dengue Cases and Population Density multiplied by Mean Index P: This plot illustrates the relationship between the number of dengue cases and the interaction of population density with the mean transmission potential (index P).

### Figure S4

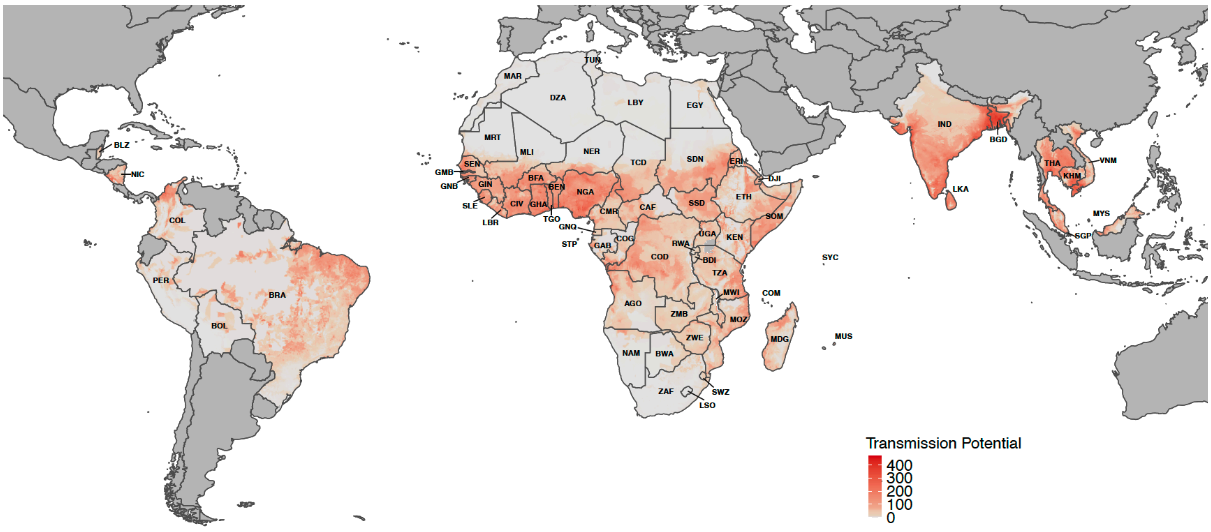


Figure S4: The figure shows the composite index of transmission suitability multiplied by Population Density (t_i), used in this study. Here we only show for the 14 countries of high incidence and the African continent, i.e., only for the origin and destination countries used in this study.

### Figure S5

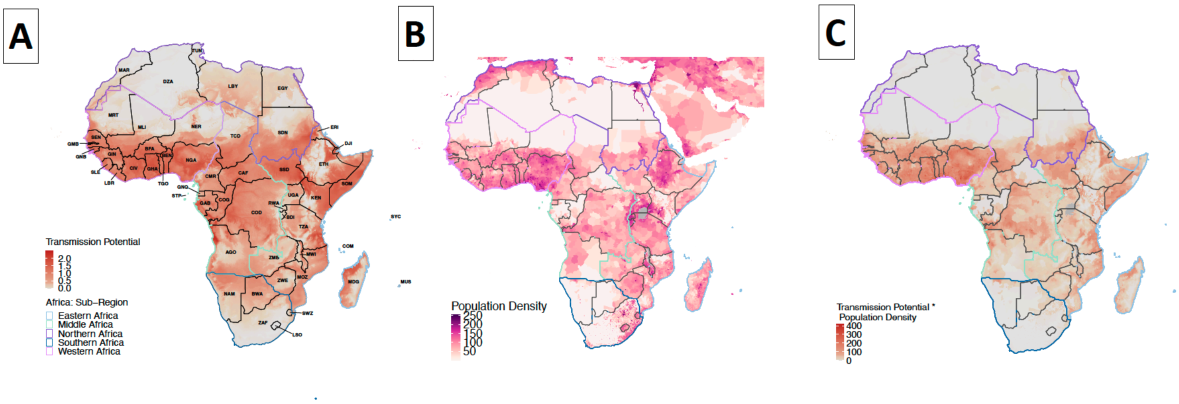


Figure S5: A) The map background uniformly represents the transmission suitability of dengue (mean index P) across Africa, with the continent divided into five distinct regions. B) The population density across Africa and C) Transmission suitability multiplied by population density for Africa. We also divide the African continent into 5 regions: Eastern (light blue) , Middle (Green), Northern (Purple), Southern (dark blue) and Western Africa (pink) which are outlined on the map.

### Figure S6

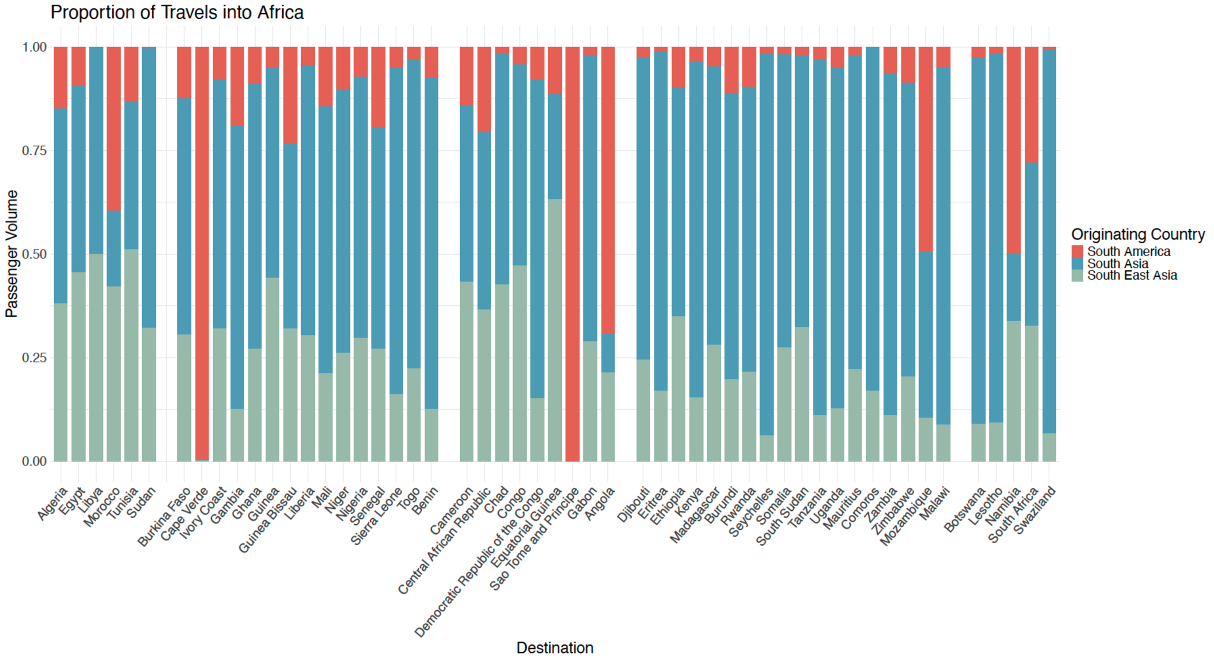


Figure S6: This figure depicts the proportion of travel volumes from Asia, Southeast Asia, and South America to African countries over the course of one year.

### Figure S7

# A

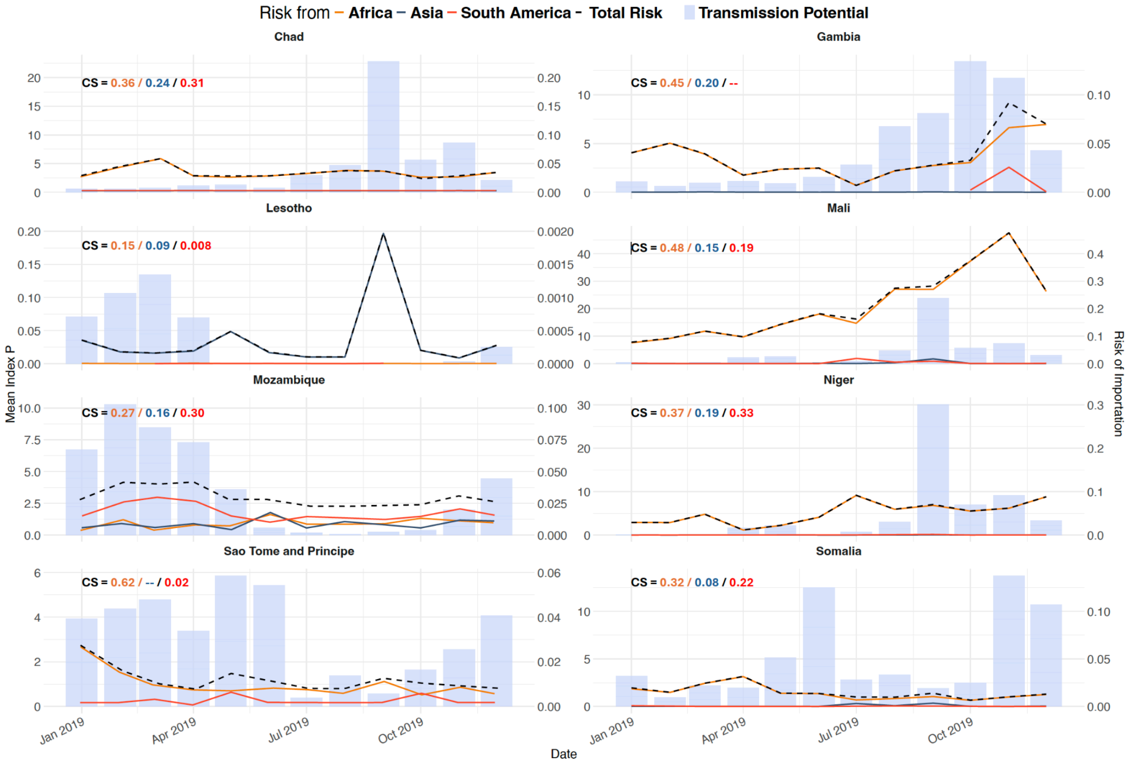

Risk from – Africa – Asia – South America - Total Risk ■ Transmission Potential
Chad
CS = 0.36 / 0.24 / 0.31
Gambia
CS = 0.45 / 0.20 / --
Lesotho
CS = 0.15 / 0.09 / 0.008
Mali
CS = 0.48 / 0.15 / 0.19
Mozambique
CS = 0.27 / 0.16 / 0.30
Niger
CS = 0.37 / 0.19 / 0.33
Sao Tome and Principe
CS = 0.62 / -- / 0.02
Somalia
CS = 0.32 / 0.08 / 0.22
Mean Index P
Risk of Importation
Jan 2019
Apr 2019
Jul 2019
Oct 2019
Date


# B

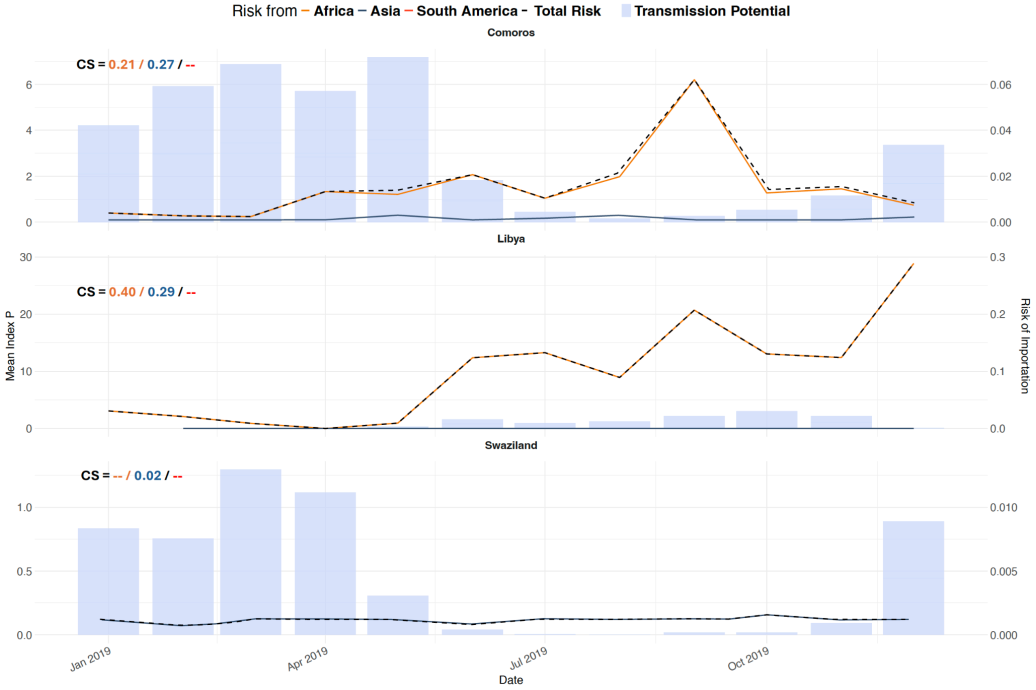

Risk from – Africa – Asia – South America - Total Risk ■ Transmission Potential
Comoros
CS = 0.21 / 0.27 / --
Libya
CS = 0.40 / 0.29 / --
Swaziland
CS = -- / 0.02 / --
Mean Index P
Risk of Importation
Jan 2019
Apr 2019
Jul 2019
Oct 2019
Date

**C**

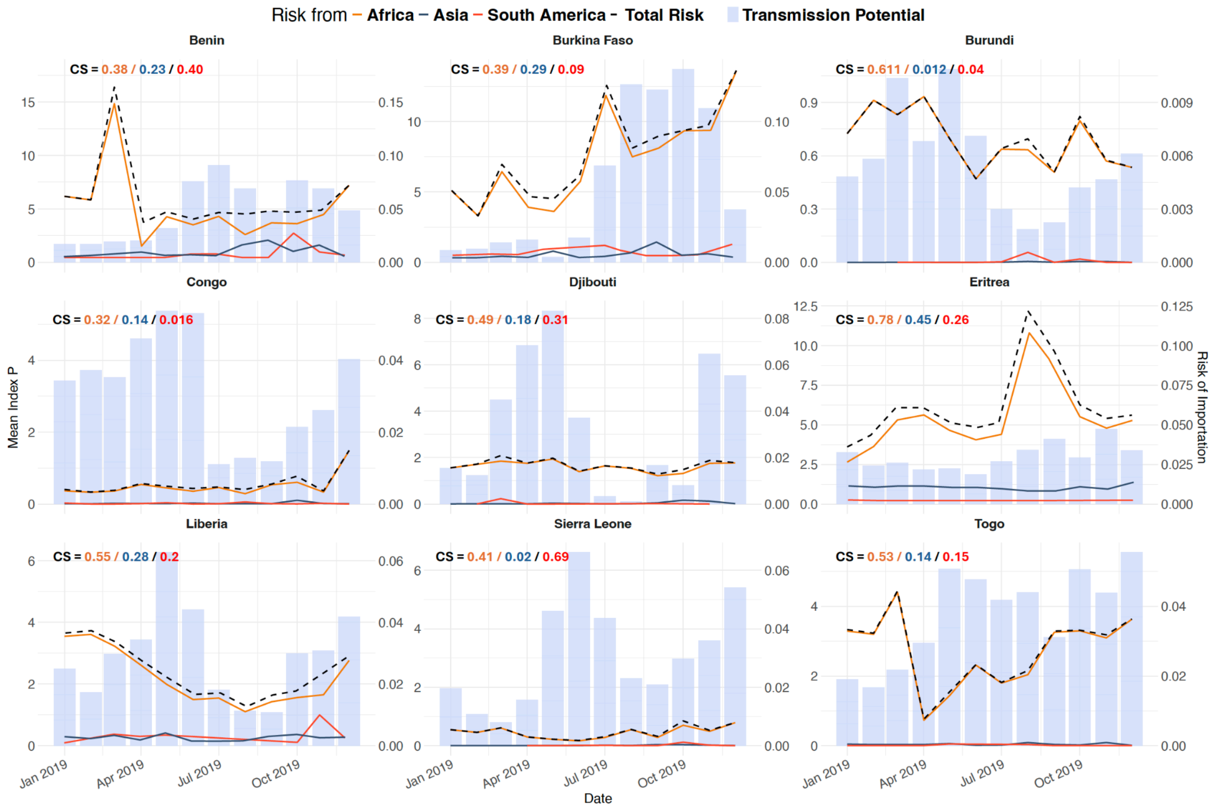
Risk from – Africa – Asia – South America - Total Risk
Transmission Potential
Benin
CS = 0.38 / 0.23 / 0.40
Burkina Faso
CS = 0.39 / 0.29 / 0.09
Burundi
CS = 0.611 / 0.012 / 0.04
Congo
CS = 0.32 / 0.14 / 0.016
Djibouti
CS = 0.49 / 0.18 / 0.31
Eritrea
CS = 0.78 / 0.45 / 0.26
Liberia
CS = 0.55 / 0.28 / 0.2
Sierra Leone
CS = 0.41 / 0.02 / 0.69
Togo
CS = 0.53 / 0.14 / 0.15
Mean Index P
Risk of Importation
Jan 2019
Apr 2019
Jul 2019
Oct 2019
Date

**D**

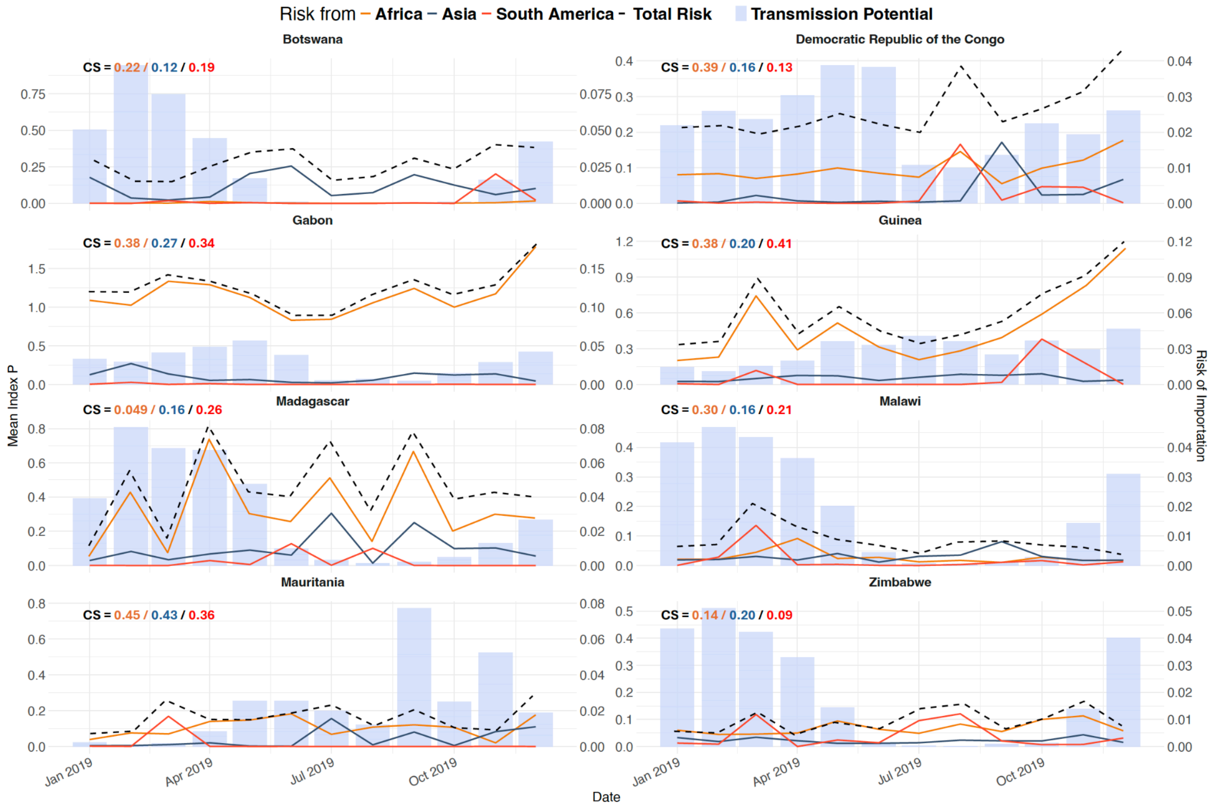
Risk from – Africa – Asia – South America - Total Risk
Transmission Potential
Botswana
CS = 0.22 / 0.12 / 0.19
Democratic Republic of the Congo
CS = 0.39 / 0.16 / 0.13
Gabon
CS = 0.38 / 0.27 / 0.34
Guinea
CS = 0.38 / 0.20 / 0.41
Madagascar
CS = 0.049 / 0.16 / 0.26
Malawi
CS = 0.30 / 0.16 / 0.21
Mauritania
CS = 0.45 / 0.43 / 0.36
Zimbabwe
CS = 0.14 / 0.20 / 0.09
Mean Index P
Risk of Importation
Jan 2019
Apr 2019
Jul 2019
Oct 2019
Date

**E**

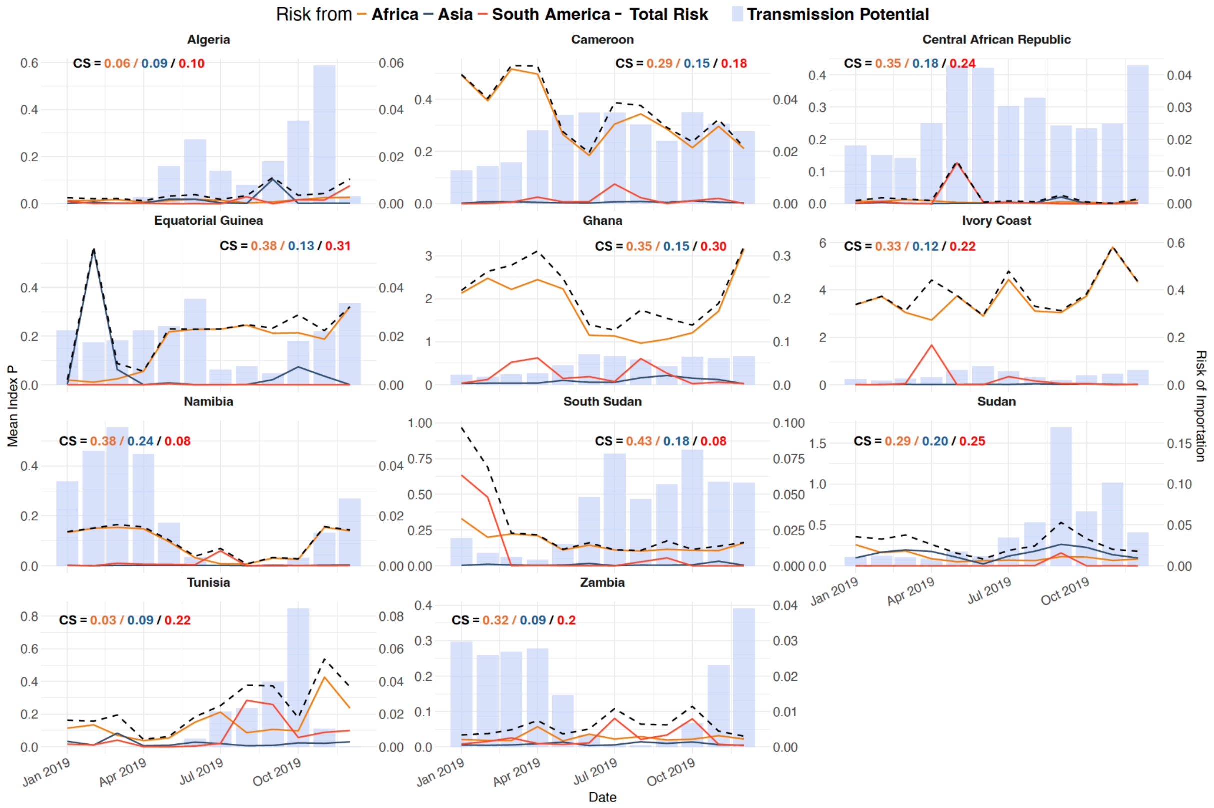


Figure S7: Time-varying risk of introduction into African countries in 2019 from Asia (blue line) and South America (red line) along with time-varying transmission suitability index nationally. The combined risk is represented by the black dotted line. Panel A) Chad, Gambia, Lesotho, Mali, Mozambique, Niger, Sao Tome and Principe, Somalia. B) Comoros, Libya, Swaziland. C) Benin, Burkina Faso, Burundi, Congo, Djibouti, Eritrea, Guinea Bissau, Liberia, Sierra Leone. D) Botswana, DRC, Gabon, Guinea, Madagascar, Malawi, Mauritania, Zimbabwe. E) Algeria, Cameroon, Central African Republic, EquatorialGuinea, Ghana, Ivory Coast, Namibia, South Sudan, Sudan and Tunisia.

### Figure S8

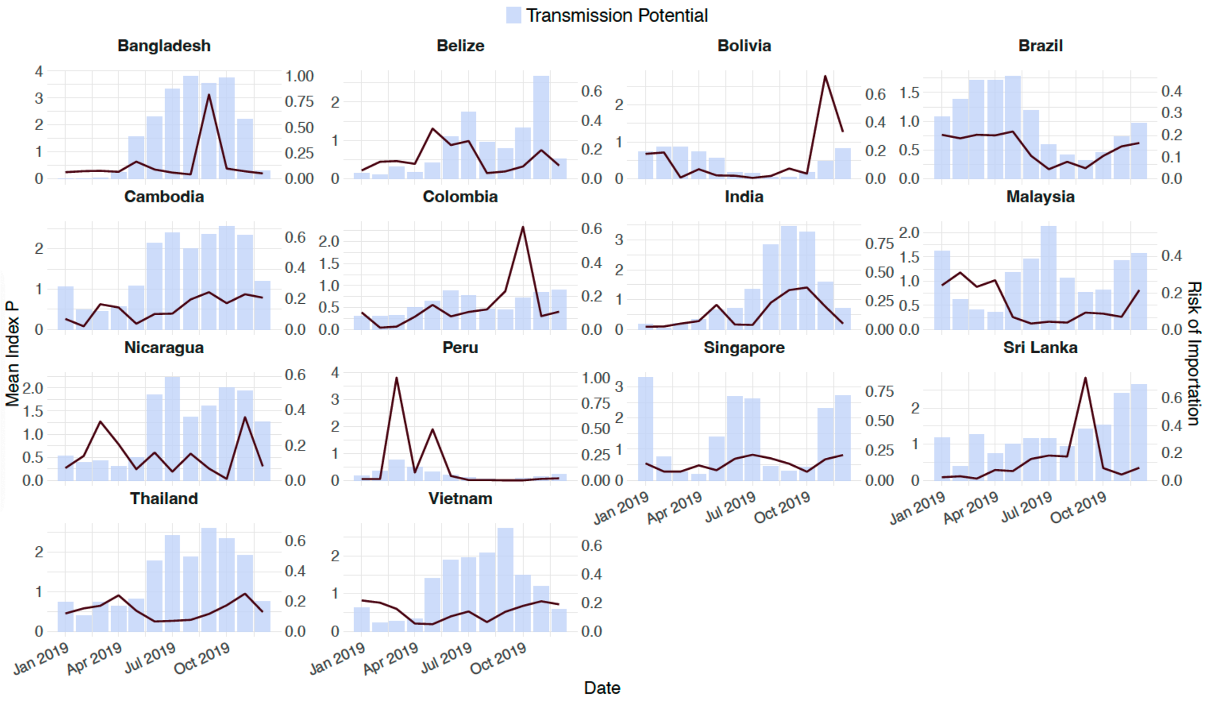


Figure S8: Risk of Exportation from countries of high incidence (n=14) considered in this study overlaid across their respective transmission suitability across the year.

#### Table S1

Table S1: Annual risk of dengue importation into various countries and the cosine similarity between the risk of importation and the local transmission suitability. The cosine similarity values highlight the level of synchrony between the timing of importation risk and transmission suitability.

| Destination Country | Africa | | Asia | | South America | |
|---|---|---|---|---|---|---|
| | Mean Annual Risk | Cosine Similarity | Mean Annual Risk | Cosine Similarity | Mean Annual Risk | Cosine Similarity |
| Algeria | 8.58793E-05 | 0.058328731 | 4.75473E-05 | 0.092625456 | 0.000128602 | 0.099100722 |
| Angola | 0.000871512 | 0.190891518 | 0.001993496 | 0.452737102 | 0.000366653 | 0.274175799 |
| Benin | 0.003930211 | 0.389007711 | 3.47791E-06 | 0.228437938 | 1.68932E-06 | 0.396225317 |
| Botswana | 3.51387E-05 | 0.215154412 | 2.13379E-05 | 0.117967166 | 2.78784E-05 | 0.194705357 |
| Burkina Faso | 0.006646177 | 0.385177331 | 4.09467E-06 | 0.286753612 | 1.88572E-06 | 0.087784552 |
| Burundi | 0.000837797 | 0.611547737 | 2.34401E-06 | 0.011767048 | 1.54542E-07 | 0.642582104 |
| Cameroon | 0.008447926 | 0.295619467 | 2.95063E-05 | 0.153247624 | 2.19541E-05 | 0.181770533 |
| Central African Republic | 0.000218083 | 0.353990993 | 3.91664E-05 | 0.177757447 | 9.86087E-06 | 0.23872773 |
| Chad | 0.000274719 | 0.369497313 | 1.53444E-06 | 0.238037186 | 2.49248E-08 | 0.316180995 |
| Comoros | 0.001294683 | 0.209741644 | 4.3464E-07 | 0.27279957 | - | - |
| Congo | 0.000586055 | 0.324397585 | 4.35204E-06 | 0.14248563 | 1.69903E-05 | 0.016785495 |
| Democratic Republic of the Congo | 0.001303986 | 0.393800624 | 8.2923E-06 | 0.162595074 | 7.14804E-06 | 0.132166652 |
| Djibouti | 0.001532898 | 0.494685428 | 7.89399E-06 | 0.187134913 | 3.04414E-08 | 0.316606702 |
| Egypt | 0.000936993 | 0.099743346 | 0.001444715 | 0.310977716 | 0.003022812 | 0.146581553 |
| Equatorial Guinea | 0.001953211 | 0.381572116 | 0.00014837 | 0.130329526 | 4.96267E-06 | 0.316211807 |
| Eritrea | 0.008478351 | 0.777500496 | 9.86055E-07 | 0.4451418 | 3.4245E-08 | 0.26141747 |
| Ethiopia | 0.005274767 | 0.150157274 | 0.000581153 | 0.00676671 | 0.000591047 | 0.194017404 |
| Gabon | 0.007230396 | 0.375102928 | 8.65829E-06 | 0.27379809 | 4.1067E-06 | 0.336726972 |
| Gambia | 0.003186559 | 0.458145605 | 8.03388E-06 | 0.201716686 | 0.005653556 | - |
| Ghana | 0.00955636 | 0.351897748 | 0.000170407 | 0.150931437 | 5.47708E-05 | 0.295709752 |
| Guinea | 0.003630093 | 0.383805206 | 1.21755E-05 | 0.19985256 | 1.44646E-07 | 0.407950221 |
| Guinea-Bissau | 0.001072629 | 0.457622528 | - | - | - | - |
| Ivory Coast | 0.020974173 | 0.325717678 | 6.57972E-05 | 0.12354295 | 0.00107263 | 0.215319706 |

| | | | | | | |
|---|---|---|---|---|---|---|
| Kenya | 0.005731245 | 0.036114895 | 0.000988367 | 0.069622909 | 0.001494575 | 0.154600402 |
| Lesotho | 3.56208E-07 | 0.15289133 | 1.02055E-05 | 0.094732527 | 3.14016E-07 | 0.0085123 |
| Liberia | 0.002223941 | 0.552593253 | 3.04256E-06 | 0.280318149 | 9.93E-08 | 0.24756365 |
| Libya | 0.015078074 | 0.398317605 | 2.5673E-07 | 0.299733067 | - | - |
| Madagascar | 0.002337154 | 0.049387505 | 2.94347E-05 | 0.168038552 | 4.71908E-07 | 0.260730525 |
| Malawi | 0.000321708 | 0.300413857 | 5.119E-06 | 0.158354803 | 1.16176E-06 | 0.211343472 |
| Mali | 0.015824618 | 0.480114221 | 3.1599E-05 | 0.151662472 | 2.32159E-06 | 0.191559015 |
| Mauritania | 0.001212314 | 0.455332323 | 1.61734E-05 | 0.438664787 | 6.37599E-07 | 0.364489628 |
| Mauritius | 9.70938E-05 | 0.130737683 | 0.001849354 | 0.437307709 | 5.49418E-05 | 0.183208942 |
| Morocco | 0.003680877 | 0.032089383 | 0.000168915 | 0.062747594 | 0.001201956 | 0.08396825 |
| Mozambique | 8.16851E-05 | 0.265098023 | 1.54534E-05 | 0.158473509 | 2.75049E-05 | 0.302516564 |
| Namibia | 0.001218896 | 0.379170333 | 4.50418E-06 | 0.243176458 | 2.5075E-05 | 0.084827259 |
| Niger | 0.006007527 | 0.373429595 | 8.00839E-06 | 0.196077917 | 2.6152E-08 | 0.332386386 |
| Nigeria | 0.002684687 | 0.175464736 | 0.000385632 | 0.129285607 | 0.003900263 | 0.166146102 |
| Rwanda | 0.001544029 | 0.534617381 | 7.87532E-05 | 0.082448877 | 7.02317E-06 | 0.178981757 |
| Sao Tome and Principe | 0.000170228 | 0.616702222 | - | - | 1.15721E-05 | 0.02254764 |
| Senegal | 0.022102646 | 0.354560297 | 2.50289E-05 | 0.078299913 | 0.00144425 | 0.090820404 |
| Sierra Leone | 0.000356306 | 0.414192321 | 4.02423E-06 | 0.020192397 | 1.28825E-08 | 0.690094408 |
| Somalia | 0.001317879 | 0.315245767 | 1.73787E-05 | 0.082469611 | 3.80101E-05 | 0.21881435 |
| South Africa | 0.005954194 | 0.123350931 | 0.001192238 | 0.102518352 | 0.003235918 | 0.100622363 |
| South Sudan | 0.001536735 | 0.43148707 | 1.00949E-05 | 0.183587161 | 0.001606043 | 0.081165463 |
| Sudan | 0.001331234 | 0.292155652 | 0.000214187 | 0.196758845 | 0.000140615 | 0.248052223 |
| Swaziland | 0 | - | 5.24909E-07 | 0.024587963 | 1.42E-08 | - |
| Tanzania | 0.004724326 | 0.252885566 | 6.78168E-05 | 0.121105075 | 3.23203E-05 | 0.161858017 |
| Togo | 0.002531176 | 0.526981151 | 4.93146E-06 | 0.141542277 | 5.21319E-06 | 0.274707511 |
| Tunisia | 0.00353866 | 0.030401063 | 9.32356E-05 | 0.085759411 | 0.00045281 | 0.223887708 |
| Uganda | 0.006595113 | 0.294246386 | 0.00036009 | 0.174638736 | 0.00022749 | 0.139245862 |
| Zambia | 0.000214647 | 0.324477152 | 3.77893E-05 | 0.088560608 | 1.53366E-05 | 0.200134605 |
| Zimbabwe | 0.00058781 | 0.146458059 | 1.07493E-05 | 0.201679279 | 1.38175E-05 | 0.094483847 |
